# Protein phosphatase 2A, complement component 4, and *APOE* genotype linked to Alzheimer’s disease using a systems biology approach

**DOI:** 10.1101/2020.11.20.20235051

**Authors:** Gyungah R. Jun, Yang You, Congcong Zhu, Gaoyuan Meng, Jaeyoon Chung, Rebecca Panitch, Junming Hu, Weiming Xia, The Alzheimer’s Disease Genetics Consortium, David A. Bennett, Tatiana M. Foroud, Li-San Wang, Jonathan L. Haines, Richard Mayeux, Margaret A. Pericak-Vance, Gerard D. Schellenberg, Rhoda Au, Kathryn L. Lunetta, Tsuneya Ikezu, Thor D. Stein, Lindsay A. Farrer

**Author notes:** To whom correspondence should be addressed Gyungah R. Jun, PhD, Department of Medicine (Biomedical Genetics), Boston University School of Medicine, 72 East Concord Street, Boston, MA 02118, USA 02118,; Lindsay A. Farrer, PhD, Department of Medicine (Biomedical Genetics), Boston University School of Medicine, 72 East Concord Street, Boston, MA 02118, USA 02118,.

## Abstract

**Background:** Recent reports suggest that the rare apolipoprotein E (*APOE*) Christchurch mutation and ε2 allele protect against Alzheimer’s disease (AD) pathology by reducing the burden of tau pathology. However, the mechanism(s) underlying the ε2 protective effect linking to tau is largely unknown.

**Methods:** The role of the ε2 allele in Alzheimer’s disease (AD) was investigated a genome-wide association study (GWAS) for AD among 2,120 ε2 carriers from the Alzheimer Disease Genetics Consortium (ADGC), and then prioritized by gene network analysis, differential gene expression analysis at tissue- and cell-levels as well as methylation profiling of CpG sites, in prefrontal cortex tissue from 761 brains of the Religious Orders Study and Memory and Aging Project (ROSMAP) and the Framingham Heart Study (FHS), Boston University Alzheimer’s Disease Center (BUADC). The levels of two catalytic subunit proteins from protein phosphatase 2A (PPP2CA and PPP2CB) were validated in prefrontal cortex area of 193 of the FHS/BUADC brains. The findings from human autopsied brains were further validated by a co-culture experiment of human isogenic *APOE* induced pluripotent stem cell (iPSC) derived neurons and astrocytes.

**Results:** Of the significantly associated loci with AD among *APOE* ε2 carriers (P<10^−6^), *PPP2CB* (P=1.1×10^−7^) was the key node in the *APOE* ε2*-*related gene network and contained the most significant CpG site (P=7.3×10^−4^) located 2,814 base pair upstream of the top-ranked GWAS variant. Among *APOE* ε3/ε4 subjects, the level of Aβ_42_ was negatively correlated with protein levels of PPP2CA (P=9.9×10^−3^) and PPP2CB (P=2.4×10^−3^), and PPP2CA level was correlated with the level of pTau231 level (P=5.3×10^−3^). Significant correlations were also observed for PPP2CB with complement 4B (C4B) protein levels (P=3.3×10^−7^) and PPP2CA with cross reactive protein (CRP) levels (P=6.4×10^−4^). C1q level was not associated with Aβ_42_, pTau231, PPP2CB, or C4B levels. We confirmed the significant correlation of *PPP2CB* expression with pTau231/tTau ratio (P=0.01) and *C4A/B* (P=2.0×10^−4^) expression observed in brain tissue in a co-culture experiment of iPSC derived neurons and astrocytes.

**Conclusion:** We demonstrated for the first time a molecular link between a tau phosphatase and the classical complement pathway, especially C4, and AD-related tau pathology.

## BACKGROUND

Alzheimer disease (AD) is a progressive neurodegenerative disorder and accounts for 60-80% of all causes of dementia. None of the prescribed medications for symptomatic treatment of AD retard or stop neuronal degeneration [1]. AD currently affects about 5.8 million Americans age 65 and older and will increase to 13.8 million by 2050 if current trends continue [1]. AD is the sixth leading cause of death in the United States and its mortality rate increased 146% between 2000 and 2018 [1]. More than 16 million caregivers provided about 18.6 billion hours of care (valued around $244 billion) to people with AD or other dementias in 2019, and total health care payments in 2020 for people age 65 and older with dementia are estimated to be $305 billion [1]. Apolipoprotein E (*APOE*) genotype is the strongest risk factor for the common form of AD that occurs after age 65 years [1]. Combinations of amino acid residues at 112 (rs429358) and 158 (rs7412) determine three common *APOE* alleles (ε2, ε3 and ε4), where ε4 increases but ε2 decreases AD risk [2, 3]. Lifetime risk of AD among female ε4 homozygotes is approximately 60% and 10 times higher than for ε2 carriers matched for sex and age [2, 3].

Neuropathological hallmarks of AD are neurofibrillary tangles consisting of decomposed microtubules and phosphorylated tau (p-tau) and neuritic plaques containing toxic beta-amyloid peptides (Aβ) [1]. Frequencies of *APOE* ε2/2, ε3/3 and ε4/4 homozygotes among non-Hispanic whites are about 0.5%, 38.2%, and 12.6%, respectively [3, 4]. Odds ratios of AD among clinically and neuropathologically confirmed ε4 and ε2 homozygotes in this group are approximately 31 and 0.13, respectively, compared to persons with the common ε3/ε3 genotype [5]. Common variants in the region of *MAPT* (the gene encoding tau protein) have been associated with AD among individuals who lack ε4 [6]. The protective effect of ε2 on tangle burden is independent from that on plaque burden and specific to AD pathology [5]. An exaggerated modifying effect of *APOE* alleles on clinical and pathological manifestations of AD was demonstrated by the recent discovery of the rare *APOE* Christchurch (APOEch) mutation on a ε3 chromosome background in a carrier of the deleterious presenilin 1 (*PSEN1*) E280A mutation that causes early onset AD typically between ages 30 and 60 among members of a large kindred with autosomal dominant AD. Notably, the E280A mutation carrier who was homozygous for the APOEch variant had hyperlipoproteinemia Type III that is typically associated with ε2 homozygosity, presented with delayed cognitive impairment in her seventies, and showed profound plaque burden but limited neurofibrillary tangle involvement by positron emission tomography (PET) imaging [7]. In fact, accumulation of tau protein (the primary constituent of tangles) measured by PET is strongly associated with memory decline and most prominent in the medial temporal lobe [8].

To evaluate the contribution of other genetic factors to the protective effect of ε2, we conducted a genome-wide association study (GWAS) for AD among ε2 carriers, applied a systems biology approach to identify biologically connected networks that contain genes showing evidence of genetic association, and demonstrated biological relevance of top-ranked genes to AD experimentally by immunoassay in brain tissue of neuropathologically confirmed AD cases and controls.

## METHODS

### Genome-wide Association Analyses

#### Subjects

The study included 34 cohorts from the Alzheimer’s Disease Genetics Consortium (ADGC) containing 14,536 subjects meeting clinical or neuropathological criteria for probable AD and 14,518 cognitively unimpaired controls for whom *APOE* genotype and genome-wide single nucleotide polymorphism (SNP) array data were available (**Table S1**) [9]. After excluding subjects younger than 65 years of age at the time of censoring (onset of symptoms for AD cases or last examination/death for controls), ε2/4 subjects, and cohorts with fewer than 10 cases and 10 controls in any of four *APOE* genotype groupings (ε2/ε2+ε2/ε3 [E2P]), ε3/ε3 [E33], ε3/ε4 [E34], ε4/ε4 [E44]), a total of 13,909 cases and 14,252 controls from 29 cohorts remained for genetic association analysis. Characteristics of these subjects are provided in **Table S2**.

#### Phenotypic Evaluation

A diagnosis of probable AD among clinically evaluated subjects in ADGC datasets was established by Diagnostic and Statistical Manual of Mental Disorders (DSM-IV) criteria, National Institute of Neurological and Communicative Disorders and Stroke (NINCDS)/Alzheimer’s Disease and Related Disorders Association (ADRDA) criteria for probable AD [10] with Clinical Diagnostic Ratings (CDRs) greater than zero [11]. Controls were cognitively unimpaired with a CDR of zero. The ADGC sample included 5,007 neuropathologically examined subjects (4,018 cases and 989 controls) [5].

#### Genotyping, quality control, and imputation

We applied quality control filters to exclude individuals with genome-wide genotype call rate <95%, discordance between reported and genetically determined sex, and individuals of non-European ancestry [9]. Single nucleotide polymorphisms (SNPs) with a call rate <95%, Hardy-Weinberg equilibrium P<10^−6^, and minor allele frequency (MAF) <0.01 were excluded. Principal components (PC) of ancestry were determined separately in each cohort using EIGENSOFT [12]. The first three PCs were included as covariates in all subsequent analyses to correct for population substructure. SNP genotype probabilities were imputed using the Haplotype Reference Consortium (HRC) reference panel and the Michigan Imputation Server (https://imputationserver.sph.umich.edu/). SNPs with MAF<0.01 and imputation quality threshold of R^2^<0.4 were excluded.

#### Genome-wide association tests

The association of AD was tested with each SNP in each *APOE* genotype group using a logistic regression model including a quantitative estimate between 0 and 2 representing the probability of the effect allele to incorporate the uncertainty of the imputation estimates and covariates to adjust for age, sex, and PCs. The MIRAGE and NIA-LOAD cohorts were analyzed using generalized estimating equation (GEE) models to account for family structure. Analyses were conducted separately in each cohort and the results were combined by meta-analysis using the inverse variance method in METAL [13]. Analyses were performed in subgroups defined by *APOE* genotype including ε2/ε2+ε2/ε3, ε3/ε3, ε3/ε4, and ε4/ε4. Analyses were performed separately for each dataset containing at least 10 cases and 10 controls and the results were combined by meta-analysis. To assess interaction between a SNP and ε2 among ε4 non-carriers (i.e., ε2/ε2 ε2/ε3 and ε3/ε3 subjects), models included a main effect for dose of ε2 coded as 0, 1, or 2 and a term for the interaction between the SNP and ε2 dosage. An analogous model was employed to assess interaction between a SNP and ε4 among ε2 non-carriers (i.e., ε3/ε3 ε3/ε4 and ε4/ε4 subjects). Odds ratios (ORs) and 95% confidence intervals (95% CIs) were computed for the SNP or *APOE*\*SNP interaction.

#### Gene network and enrichment analysis

We conducted gene network analyses stratified by *APOE* genotype that were seeded with genes containing SNPs that were significantly associated with AD (P<10^−5^) in corresponding *APOE* genotype groups using the using the Ingenuity Pathway Analysis (IPA, QIAGEN, Redwood, CA) and data derived experimentally in animal and cell models to identify statistically over-represented biological functions or pathways. We selected networks with a score >10 that contained more than five seed genes for enrichment analysis. We performed enrichment tests on those *APOE* related networks with the gene sets for neuropathological traits including neuritic plaques (NP) and neurofibrillary tangles (NFT) using the Fisher’s exact test implemented in R. The genes in those neuropathological gene sets were defined by the locations of SNPs with P≤0.001 in the previous GWAS for NP measured according to the CERAD score and NFT measured by Braak stages [14]. To identify the shared biological functions and pathways among the *APOE*-related networks, we conducted pathway analysis using IPA using the same parameters used in network analysis.

### Analysis of Top-Ranked Genes Using Brain Omics Data

#### Source of autopsied brains, phenotypic definitions, and gene expression levels

RNA sequence data including gene expression information derived from dorsolateral prefrontal cortex area tissue donated by 627 participants (380 autopsy-confirmed AD cases and 247 controls) in the Religious Orders Study and Memory and Aging Project (ROSMAP) [15, 16] were obtained from the Synapse website (http://www.synapse.org). We also obtained analogous data derived from the same brain region donated by 208 participants (64 autopsy-confirmed AD cases and 129 controls) in the Framingham Heart Study (FHS) and Boston University Alzheimer’s Disease Center (BUADC). The National Institute of Aging-Reagan criteria were used to diagnose AD, which included intermediate or high probability [17]. A diagnosis of definite AD among subjects in both ROSMAP and FHS/BUADC cohorts was established according to the National Institute of Aging (NIA) Reagan criteria [18-20]. Consortium to Establish a Registry for Alzheimer Disease (CERAD) semi-quantitative neuritic plaque scores were derived by coding 1 for definite, 2 for probable, 3 for possible, and 4 for none [21]. Braak stages reflecting the spatial distribution of neurofibrillary tangles were coded as 0 for none, I-II (transentorhinal and entorhinal cortex), III-IV (hippocampal and neighboring limbic areas), and V-VI (neocortical areas) [19]. The sample included 403 AD cases and 358 controls in the combined cohorts.

#### Gene expression analysis

We selected 111 genes from the APOE genotype group networks containing at least one SNP with P<10^−5^ from the corresponding APOE genotype group GWAS. Gene expression levels were compared between AD and control brains using linear regression models including log-transformed normalized expression values from bulk RNA sequencing (bulk RNA-seq) data and covariates. For cell-type specific expression profiling and enrichment tests, we also downloaded single-nucleus RNA sequencing (snRNA-seq) data derived from the prefrontal cortex (Broadman area 10) of 24 AD and 24 control brains in the ROSMAP cohort from the Synapse website. Details of quality control, processing, and normalization of de novo bulk RNA-seq and snRNA-seq data as well as gene expression analysis were provided in **Supplementary Note**. Covariates for ROSMAP included age at death, sex, RNA integrity number (RIN), batch effect, and education level. Models for analysis of FHS/BUADC data included age at death sex, RIN, post-mortem interval (PMI), and batch number. Analyses were conducted in the total sample and separately in *APOE* genotype groups (ε2/3, ε3/3, and ε3/4) in each cohort. There were insufficient numbers of ε2/2 or ε4/4 subjects for analyses within these genotype groups. Results from each cohort were combined by meta-analysis as described above using the number of AD cases as a weight and log2 of fold change (Log2FC) as a direction. Association of gene expression with plaque score and Braak stage was evaluated in each cohort using a GLM model including the covariates listed above, and the results from the two datasets were combined by meta-analysis using the inverse variance method in METAL [13]. The Bonferroni corrected significance threshold for these comparisons was P=4.5×10^−4^.

Single-nucleus RNA sequencing (snRNA-seq) data derived from the prefrontal cortex (Broadman area 10) of 24 AD and 24 control brains in the ROSMAP cohort were obtained from the Synapse website. Quality control and data processing of these data are described in **Supplementary Note**. Normalized gene expression measures from each cell were scaled by 10,000 multiplied by the total library size and then log transformed using the ScaleData function in R. We conducted principle component analysis of cell type specific expression levels using data for previously established marker genes for each brain cell type [22] and t-distributed Stochastic Neighbor Embedding (tSNE) method [23]. This analysis yielded eight cell-type specific clusters including astrocytes (Ast), endothelial cells (End), excitatory neurons (ExN), inhibitory neurons (InN), microglia (Mic), oligodendrocytes (Oli), oligodendrocyte progenitor cells (OPC), and pericytes. Endothelial cells and pericytes were excluded from further analysis due to their low proportion. The percentage of expressed cells for each cell type in each sample was visualized using bar and violin plots. Distributions of expressed cells for top ranked genes were evaluated using the tSNE program in R. Log transformed fold changes (Log2FC) in expression between AD and control samples were evaluated for each of the top-ranked genes within each cell type using the Wilcoxon rank-sum test. False discovery rate (FDR) p-values were adjusted using the Benjamin-Hochberg procedure implemented in R [24].

#### Methylation data analysis

We obtained methylation array data obtained for 697 ROSMAP subjects from the Synapse website [16]. We assessed the percentage of methylation of all CpG sites spanning the region encompassed by 10,000 base pairs upstream and downstream of each gene. Association of the degree of methylation at each CpG site with neuropathologically defined AD, and with rank-transformed quantitative measures of plaques and tangles, was evaluated in regression models including covariates for sex, age, and batch effect.

### Validation of the Top-Ranked Genes in Human Brain Tissue

Functional relevance of PP2A subunits encoded by *PPP2CB* and *PPP2CA* to AD was evaluated by immunoassays of AD-related biomarkers included Aβ_42_, phosphorylated tau 181 (pTau181), phosphorylated tau 231 (pTau231), and total tau (tTau) in brain tissue donated by 224 FHS and BUADC participants. We also assessed classical complement proteins C1q, C4A, and C4B, as well as CRP, because recent studies demonstrated ApoE isoforms bind C1q protein and modulate classical complement-dependent synapse loss and neuroinflammation [25], plasma CRP levels modulate *APOE* genotype-dependent onset of AD [26].

#### Immunoassay measurement

Frozen tissue from the dorsolateral prefrontal cortex (Brodmann area 8/9) of the FHS/BUADC brains was weighed and placed on dry ice (n=224). Freshly prepared, ice cold 5M Guanidine Hydrochloride in Tris-buffered saline (20 mM Tris-HCl, 150 mM NaCl, pH 7.4) containing 1:100 Halt protease inhibitor cocktail (Thermo Fischer Scientific, Waltham, MA) and 1:100 Phosphatase inhibitor cocktail 2 & 3 (Sigma-Aldrich, St. Louis, MO) was added to the brain tissue at 5:1 (5M Guanidine Hydrochloride volume (ml):brain wet weight (g)) and homogenized with Qiagen Tissue Lyser LT at 50Hz for five min. The homogenate was then mixed (regular rocker) overnight at room temperature. The lysate was diluted with 1% Blocker A (Meso Scale Discovery (MSD), Rockville, Maryland, #R93BA-4) in wash buffer according to specific immunoassays: 1:300 for total tau, pTau231 (MSD #K15121D-2), and pTau181 (MSD #K250QND), and 1:4000 for Aβ42 (MSD #K15200E-2). Samples were subsequently centrifuged at 17,000g and 4°C for 15 minutes, after which the supernatant was applied to the immunoassays. To capture tau phosphorylated at Thr residue 181, antibody AT270 was used and the detecting antibody was the biotinylated HT7 that recognizes residue 159–163 of tau (Thermo Scientific, Rockford, IL). Internal calibrators of p-tau and tau were used (MSD). Measurements were performed using the multi-detection SPECTOR 6000 Imager (MSD).

For C4A (Novus Biologicals, NBP2-70043), C4B (Novus Biologicals, NBP2-70046, PPP2CA, and PPP2CB (MyBioSource, MBS9324930), ice cold PBS buffer (Gibco, ref#10010-023) was added to the brain tissue at 5:1 (PBS in ml vs brain wet weight in gram), and homogenized with Qiagen Tissue Lyser LT at 50Hz for five min. The homogenate was centrifuged at 17,000g and 4°C for 15 minutes, then the supernatant was aliquoted and stored at −80°C. Lysate was diluted 1:200 (C4A) or left undiluted (C4B and PPP2CB) and applied to ELISA assays according to the manufacturer’s protocol. For PPP2CA, lysate was diluted 1:10 by using sample diluent buffer, centrifuged at 17,000g and 4°C for 15 minutes, then the supernatant was applied to the assay (Novatein Biosciences, NR-E10765). Absorbance was measured at 450 nm using Biotek Synergy HT microplate reader. All immunoassay measures from the FHS/BUADC brains were adjusted for age at death and sex, and then the residuals of the measures were rank-transformed. The rank-transformed measures were used as quantitative outcomes in subsequent association analysis.

#### Association analysis methods

Association of log2-transformed gene expression values with rank-transformed immunoassay measures of Aβ42, pTau181/tTau ratio, pTau231/tTau ratio, PPP2CB, PP2A, C1q, C4A, C4B, and CRP was evaluated using linear regression models including covariates for age at death and sex. Analyses were conducted in the total sample as well as in subgroups of subjects with and without the *APOE* ε4 allele.

### Validation of *PPP2CB* and *C4A/B* Expression in Human iPSC Cells

#### Human induced pluripotent stem cell (iPSC) culture

The human parental iPSC line for *APOE* ε4/ε4, isogenic iPSC lines for *APOE* ε2/ε2 or *APOE* ε3/ε3, and an *APOE* knock-out (NO) iPSC line were purchased from ALSTEM (ALSTEM, INC., Richmond, CA). Pluripotency of the iPSC lines was fully characterized using the Human Pluripotent Stem Cell Marker Antibody Panel (anti-OCT3/4, anti-NANOG, anti-Alkaline Phosphatase, anti-SSEA-4; R&D Systems) according to the manufacturer’s instructions, and *APOE* genotype was confirmed by sequencing. iPSCs were maintained in feeder-free conditions coated with hESC-Qualified Matrigel (Corning) in complete mTeSR1 (StemCell Technologies) in a humidified incubator (5% CO_2_, 37°C). iPSCs were fed fresh media daily and passaged every 5 days. All cell lines used have been authenticated and tested negative for mycoplasma.

#### Differentiation of isogenic iPSCs into neurons (iNeurons) by NGN2 overexpression

We performed rapid induction of iPSCs into human excitatory neurons via a doxycycline-inducible neurogenin2 (NGN2) system as previously described [27, 28]. The lentiviral DNA pLV-TetO-hNGN2-eGFP-Puro was a gift from Kristen Brennand (Addgene plasmid #79823) and FUdeltaGW-rtTA was a gift from Konrad Hochedlinger (Addgene plasmid #19780). Lentiviruses were produced in HEK293T cells (ATCC) as previously described [28]. On day −1, iPSCs were dissociated into single cells with Gentle Cell Dissociation Reagent (StemCell Technologies) and plated at a density of 50,000 cells/cm^2^. Lentiviruses were added in fresh mTeSR1 medium on day 0. To induce NGN2 expression, doxycycline was added on day 1 (D1) at a final concentration of 2 µg/mL in fresh media consisting of KnockOut DMEM, KnockOut Serum Replacement, MEM-NEAA, Glutamax, and β-mercaptoethanol (all from Invitrogen). On D2, puromycin was added at 5 µg/mL for selection. On D4, cells were replated on poly-L-ornithine (10 µg/mL; Sigma-Aldrich) and mouse laminin (5 µg/mL; Invitrogen) co-coated 6-well plates (1.5-2 × 10^5^ cells/well) in media consisting of Neurobasal, B27, Glutamax, 20% dextrose, MEM-NEAA (all from Gibco) supplemented with 10 ng/mL BDNF, GDNF and CNTF (all from PeproTech). Half of the culture media was replaced every three days with fresh complete media. On D24, cells were collected for subsequent experimental assays. iNeurons were characterized by immunocytochemistry staining of the specific neuronal markers.

#### Differentiation of isogenic iPSCs into astrocytes (iAstrocytes)

Human iPSCs were differentiated to neural progenitor cells (NPCs) and subsequently to astrocytes as previously described with minor modifications [29]. Briefly, human iPSCs were plated at 10,000 cells per microwell in an AggreWell 800 24-well plate (StemCell Technologies) and differentiated to NPCs using the STEMdiff SMADi Neural Induction Kit (StemCell Technologies). On D5, the embryoid bodies (EBs) were harvested and replated onto Matrigel pre-coated 6-well plates in neural induction medium with SMAD inhibitors. Neural rosettes were selected at D12 by Rosette Selection Reagent (StemCell Technologies) and patterned to NPCs with neural induction medium and SMAD inhibitors. NPCs were characterized immunocytochemically for neural progenitor cell markers. Further, single cells of passaged NPCs (15,000 cells/cm^2^) were dissociated and differentiated to astrocytes in astrocyte medium (ScienCell) on Matrigel coated culture wares as preciously described [29]. Cells were continually cultured and harvested as astrocytes after 6 times passages (around D30). Astrocytes were validated immunocytochemically for the astrocyte-specific markers and used for co-culture experiments.

#### Co-culture system of human iPSC-derived neurons and astrocytes

Human iPSC-derived astrocytes after day 54 of differentiation were plated on poly-L-ornithine/laminin-coated hanging cell culture inserts with 1μm diameter pores used in a 6-well plate (Millipore). The differentiated neurons at day 15 were co-cultured with the inserts containing astrocytes with the corresponding isogenic *APOE* genotypes in astrocytic medium containing dialyzed FBS (Invitrogen) as previously described with some modifications [30]. After nine days upon co-culture, neurons were collected for subsequent experimental assays.

#### Immunocytochemistry (ICC)

Human iPSCs and differentiated central nervous system (CNS) cells were washed three times with 1×DPBS and fixed in 4% paraformaldehyde at 37°C for 15 min. Cells were permeabilized and blocked in blocking solution [1×PBS, 5% bovine serum albumin (BSA), 5% goat or donkey serum, 1% Triton X-100] at room temperature for 1 hr. Cells were then labelled overnight at 4°C and the primary antibodies used for hiPSCs were anti-OCT3/4, anti-NANOG, anti-Alkaline Phosphatase, and anti-SSEA-4 (all1:100, R&D Systems, SC008); for neurons were anti-MAP2 (1:1000, Millipore, AB5622), and anti-SYN1 (1:1000, Cell Signaling Technology, 5297S); for neural progenitor cells were anti-SOX2 (1:1000, R&D, AF1828), anti-PAX6 (1:1000, R&D, AF8150) and anti-NESTIN (1:1000, R&D, MAB1259); for astrocytes were anti-GFAP (1:1000, Agilent, Z033429), anti-S100β (1:1000, Sigma, AMAB91038) and anti-Vimentin (1:1000, Cell Signaling Technology, 5741T). Alexa Fluor® secondary antibodies (Life Technologies) were conjugated to their target species at room temperature for 1 hr in blocking buffer. DAPI (4’,6-diamidino-2-phenylindole, 1:2500, Thermo Fisher Scientific, D1306) was used to label nuclei. Images were acquired using a Leica SP8 Confocal Microscope.

#### Total RNA isolation and quantitative RT-PCR

Total RNA was extracted from cells using Qiazol (Qiagen) and miRNeasy kit (Qiagen) according to the manufacturer’s protocol. RNA was quantified using NanoDrop (Thermo Fisher Scientific), and RNA quality was assessed by 260/280 and 260/230 ratios. For conventional quantitative reverse transcription polymerase chain reaction (qRT-PCR), total RNA (150 ng) with specific mRNA probes were used after reverse transcription reaction according to the manufacturer (SuperScript Vilo IV Kit, Invitrogen). All mRNA amplifications were performed using commercially available FAM-labeled Taqman probes with Taqman Fast Advanced Master Mix (Applied Biosystems/Thermo Fisher Scientific). The following Taqman probes were used: Hs00416393_g1-C4A/B, Hs00602137_m1-PPP2CB, Hs99999905_m1-GAPDH. All qRT-PCRs were performed in duplicate, and the data were presented as relative expression compared to GAPDH (reference gene) as mean ± standard error (SEM). Real-time PCR reactions were performed using the 7900HT PCR system (Applied Biosystems). No RT control reactions were run to confirm successful DNA contamination removal.

#### Enzyme-linked immunosorbent assay (ELISA)

Induced *APOE* KO, ε2/ε2, ε3/ε3, and ε4/ε4 neurons were collected and lysated in RIPA buffer (Thermo Fisher Scientific) supplemented with 1% Triton-X100 and Pierce HALT inhibitor (Thermo Fisher Scientific) for analysis of cellular protein amounts by the bicinchoninic acid (BCA) assay (Thermo Fisher Scientific) and tTau and pTau levels by ELISA. Commercially available kits (tTau: #KHB0042, pTau181: #KHO0631, pTau231: #KHB8051, Thermo Fisher Scientific; Aβ40: DAB140B, Aβ42: DAB142, R&D Systems) were used to assess tTau, pTau and Aβ levels according to manufacturer’s instructions. Medium from neuronal cultures was harvested for human APOE measurement by Human Apolipoprotein E ELISA Kit (Abcam, AB108813). All measurements and data analyses were performed blind to *APOE* genotype. We performed six replicates within each iPSC line for the quantitative experiments.

#### Statistical analysis

Graphical data were analyzed using GraphPad Prism version 6.0. Data were presented as mean ± SEM and tested for normality by Kolmogorov-Smirnov test. Comparisons among four groups were analyzed by ordinary one-way ANOVA with Tukey’s multiple test corrections if data passed the normality test, and otherwise by a nonparametric test (Kruskal-Wallis test) with Dunn’s multiple test correction. Statistical analyses including the calculation of Pearson correlation coefficients were performed using the GraphPad Prism software.

## RESULTS

### Novel *APOE* genotype-specific associations with AD

**Fig. S1** illustrates our approach to identify genes that may contribute to the *APOE* ε2 protective effect on AD. We conducted a genome-wide association study (GWAS) to search for *APOE* genotype-specific associations with AD in a data set comprised of 34 cohorts assembled by the Alzheimer’s Disease Genetics Consortium (ADGC) that contained genome-wide single nucleotide polymorphism (SNP) information for 13,909 AD cases and 14,206 cognitively normal controls (**Tables S1 and S2**). There was little evidence of genomic inflation (lamda>1) except in strata with relatively small sample sizes, especially the ε2 group including persons with *APOE* genotypes ε2/ε2and ε2/ε3 (**Fig. S2a**). Due to small number of ε2 carriers, we further tested additive models including a term for the interaction between the number of ε2 alleles and a SNP (**Fig. S2b**). The top-rankedε2-related findings for main and interaction effects (**Table 1, Fig. S3a-b, Fig. S4, and Table S3**) included genome-wide significant (GWS) associations (P<5×10^−8^) with *LOC105374313* among ε2 subjects and for interactions of ε2 with *ETV1* and *CGNL1*.The association of AD with *PPP2CB* SNP rs117296832 was nearly GWS (P=1.1×10^−7^). Many significant findings were observed among subjects with other *APOE* genotypes (**Fig. S3c-f and Table S4)** including novel GWS associations with *TRIM37* among ε3/ε3 subjects (**Fig. S5**) and with *MIR3681HG-TRIB2, KLHL29, CTNNA2*, and *ZNF443* among ε4/ε4 subjects (**Fig. S6**). There was also a GWS association for the interaction of ε4 with *NKX2-3* (**Fig. S7**).

**Table 1.** Novel significant associations (P<10^−6^) for Alzheimer disease risk among *APO*E ε2 carriers

| SNP | Locus | CH | MA | MAF | SNP Main Effect |  |  | Interaction with ε2 |  |  |
| --- | --- | --- | --- | --- | --- | --- | --- | --- | --- | --- |
|  |  |  |  |  | OR | 95% CI | P-value | OR | 95% CI | P-value |
| Genome-wide significant signals: |  |  |  |  |  |  |  |  |  |  |
| rs17239735 | CGNL1 | 15 | C | 0.19 | 1.72 | 1.37 - 2.16 | 3.8x10<sup>-6</sup> | 1.83 | 1.49 - 2.24 | 4.8x10<sup>-9</sup> |
| rs77454027 | ETV1 | 7 | C | 0.02 | 3.13 | 1.58 - 6.19 | 1.0x10<sup>-3</sup> | 5.38 | 2.98 - 9.73 | 2.5x10<sup>-8</sup> |
| rs77907104 | LOC105374313 | 3 | C | 0.02 | 8.67 | 4.29 - 17.5 | 1.7x10<sup>-9</sup> | NA | NA | NA |
| Suggestive signals: |  |  |  |  |  |  |  |  |  |  |
| rs117296832 | PPP2CB | 8 | A | 0.03 | 3.94 | 2.37 - 6.54 | 1.1x10<sup>-7</sup> | 2.46 | 1.56 - 3.86 | 9.8x10<sup>-5</sup> |
| rs76084405 | STAT5B | 17 | A | 0.03 | 4.84 | 2.68 - 8.72 | 1.6x10<sup>-7</sup> | 4.27 | 2.33 - 7.80 | 2.5x10<sup>-6</sup> |
| rs78802006 | KDM4C | 9 | C | 0.04 | 3.79 | 2.25 - 6.40 | 6.0x10<sup>-7</sup> | 2.66 | 1.53 - 4.63 | 5.6x10<sup>-4</sup> |
| rs17038845 | RIC8B | 12 | A | 0.03 | 3.55 | 2.16 - 5.86 | 6.7x10<sup>-7</sup> | 2.55 | 1.58 - 4.13 | 1.3x10<sup>-4</sup> |
| rs57056064 | RBFOX1 | 16 | C | 0.06 | 2.34 | 1.67 - 3.28 | 7.7x10<sup>-7</sup> | 1.88 | 1.37 - 2.57 | 7.8x10<sup>-5</sup> |
| rs75737704 | FBXW7 | 4 | A | 0.02 | 2.05 | 0.94 - 4.49 | 0.07 | 5.85 | 3.06 - 11.2 | 9.8x10<sup>-8</sup> |
| rs77786537 | RIPOR2 | 6 | G | 0.11 | 1.97 | 1.51 - 2.56 | 5.8x10<sup>-7</sup> | 1.94 | 1.52 - 2.49 | 1.1x10<sup>-7</sup> |
| rs9478555 | CNKSR3 | 6 | A | 0.17 | 1.53 | 1.22 - 1.92 | 2.4x10<sup>-4</sup> | 1.70 | 1.38 - 2.08 | 4.6x10<sup>-7</sup> |
| rs62239974 | CTDSPL | 3 | A | 0.05 | 2.64 | 1.70 - 4.09 | 1.4x10<sup>-5</sup> | 2.70 | 1.82 – 4.00 | 7.0x10<sup>-7</sup> |
| rs118165364 | LINC01029 | 18 | G | 0.02 | 2.15 | 1.02 - 4.55 | 0.05 | 4.52 | 2.49 - 8.22 | 7.4x10<sup>-7</sup> |
| rs77005801 | HTR1A | 5 | G | 0.04 | 2.86 | 1.79 - 4.56 | 1.1x10<sup>-5</sup> | 2.89 | 1.89 - 4.40 | 8.5x10<sup>-7</sup> |
| rs6485360 | HNRNPKP3 | 11 | A | 0.03 | 2.70 | 1.72 - 4.24 | 1.6x10<sup>-5</sup> | 2.93 | 1.91 - 4.51 | 9.6x10<sup>-7</sup> |

CH: chromosome, MA: minor allele, MAF: minor allele frequency, OR: odds ratio, 95% CI: 95% confidence interval

Gene network analyses for each *APOE* genotype group that were seeded with genes containing AD-associated SNPs (P<10^−5^) in the respective groups (**Tables S5 and S6**) identified eleven biological networks including 111 seed genes from *APOE* genotype groups were identified (**Table S6**). Scrutiny of the top-ranked networks (P<10^−4^) containing more than 5 seed genes revealed that the E2P.1, E34.1, and E44.1 networks were significantly enriched with genes previously associated with density of neuritic plaques measured by the CERAD score and neurofibrillary tangles (NFT) measured by Braak staging (**Table S7**). *PPP2CB* was identified as a hub gene in the ε2-related network with significant enrichment with plaques and tangles, while the network contained nine suggestive GWAS genes in ε2 carriers, *ARHGAP32, DNTTIP1, ITGA1, KDM4C, PMEPA1, PTPRT, STAT3, TP63*, and *UBE2C* (**Fig. S8**). The ε2-related networks collectively contained several AD-related pathways including apoptosis and several pathways involving the immune system and mitotic spindle checkpoint pathway. The E34.1 network was enriched for genes linked to WNT signaling, de-ubiquitination, and regulation of the complement cascade pathway. Genes in the E44.1 network are involved in eight pathways including ones related to nervous system development and infectious disease (**Table S7**).

### Brain transcriptomic and methylation evidence for ε2-related genes in AD

We investigated differential expression in prefrontal cortex region in neuropathologically examined brains from 403 AD cases and 358 controls (**Table S8**) for three genes showing GWS evidence of association with AD among ε2 subjects or through interaction with ε2 (*CGNL1, ETV1*, and *LOC105374313*) and 35 genes in the ε2 networks showing suggestive evidence (P<10^−5^) of association with AD (**Table S6**). Nominally significant differences in expression between AD cases and controls were observed for half (19/38) of these genes, among which upregulated expression in AD cases was transcriptome-wide significant (P<10^−6^) for *CGNL1, ECE1*, and *DNAJC1* (**Table S9**). Expression of ten of the 19 genes was also associated with plaque and/or tangle density. Twenty-seven of the 38 ε2-related genes were differentially expressed between AD and control excitatory neurons, and 12 of them were differentially expressed in inhibitory neurons (adjusted P<0.05) (**Table S10**). Evaluation of gene expression in brain cells showed that *PPP2CB* is highly expressed in excitatory neurons in both AD cases and controls, but predominantly in inhibitory neurons from controls (**Figs. S9a-c**). *PPP2CB* expression was also evident in some nuclei from other cell types (**Fig. S9d**). Expression of *APOE* (**Table S11**) was a notable exception to these trends because it was higher in excitatory neurons (P=8.6×10^−6^), inhibitory neurons (P=5.4×10^−4^), and microglia (P=0.0016), but lower in astrocytes (P=7.3×10^−13^), oligodendrocytes (P=0.0036), and OPCs (P=0.0098).

The degree of methylation at CpG sites within 10 kb from the top ranked GWAS SNPs in 11 of the 38 ε2-related genes was nominally associated with autopsy-confirmed AD diagnosis and/or severity of plaques and tangles in ROSMAP subjects (**Table S12**). Associations of methylation at CpG sites near *STAT5B* with AD status (P=3.3×10^−4^) and adjacent to *PPP2CB* with plaques (P=7.3×10^−4^) were significant after multiple test correction (**Table 2**). The methylated CpG site in *PPP2CB* (cg23693487) is located 2,814 base pairs upstream of rs117296832 which was the most significantly associated SNP in the gene with AD (**Tables 1 and 2**).

**Table 2.**
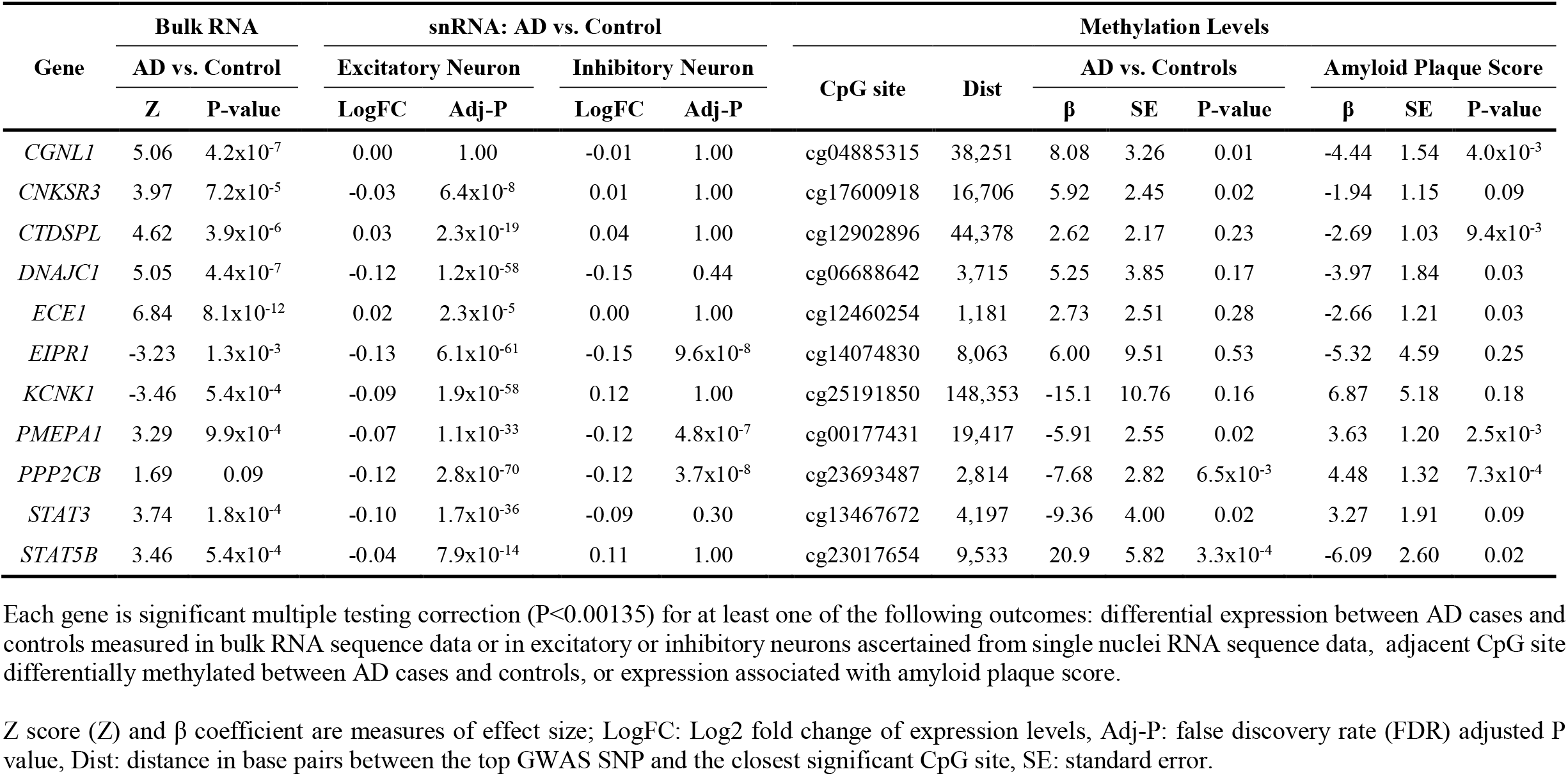
Biological role for AD-associated genes in the *APO*E ε2gene networks supported by multiple brain omics data

Eleven of the 38 *APO*E ε2-related genes were supported by multiple types of omics data derived from brain (**Table 2**). Expression of only two genes, *PMEPA1* and *PPP2CB*, was significantly lower in AD cases than controls in excitatory (adjusted P=1.1×10^−33^ and adjusted P=2.8×10^−70^, respectively) and inhibitory (adjusted P=4.8×10^−7^ and adjusted P=3.7×10^−8^, respectively) neurons, and associated with differential methylation of an adjacent CpG site between AD cases and controls (P=0.02 and P=0.0065, respectively) and density of amyloid plaques (P=0.0025 and P=7.3×10^−4^, respectively).

### Strong correlation between protein levels of PPP2CB and C4B in brain

We followed up 11 top ranked ε2-related genes supported from multiple evidence of brain omics data (**Table 2**) and 28 established late-onset AD genes (**Table S11**) with immunoassay measures of Aβ_42_ and ratio of phosphorylated tau at sites 181 (pTau181) and 231 (pTau231) to total tau (tTau), as well as protein levels of PPP2CA, PPP2CB, C4A, C4B, C1q, and CRP derived from the prefrontal cortex area of 224 FHS/BUADC brains (**Table 3**). PPP2CB level was significantly lower (P=0.008) but the Aβ_42_ level was significantly higher (P=0.005) among ε4 carriers compared to subjects lacking ε4 (**Table 3 and Fig. S10a**). Levels of other proteins including PPP2CA were not different between ε4 carriers and non-carriers (**Table 3**). As expected, protein levels of PPP2CA and PPP2CB were significantly correlated (P=2×10^−7^) and this pattern was very similar among ε4 non-carriers (**Table 4 and Fig. S10b**). Both PPP2CA and PPP2CB levels were negatively correlated with Aβ_42_ level among ε3/ε4 subjects (P<0.01), although the PPP2CA result was borderline significant after correcting for testing 11 proteins (**Table 4 and Fig. S10c**). The levels of PPP2CA and PPP2CB were inversely associated with the Aβ_42_ level (P<0.01), while the level of PPP2CA, but not PPP2CB, was positively associated with the pTau231 level among ε3/ε4 subjects (P=0.0053). These results suggest that in the presence of *APOE* ε4 both PPP2CA and PPP2CB may have a role in processing Aβ, and PPP2CA may have a major role in tau phosphorylation. Comparison of levels for PPP2CA and PPP2CB with those for the complement proteins revealed significant association of PPP2CB level with C4B in the entire sample (P=3.3×10^−7^), a pattern which was evident among subjects with and without ε4 (**Table 4 and Fig. S10d**). In contrast, levels of both PPP2CA and PPP2CB were uncorrelated with C4A level (**Table 4**). We also observed significant association between PPP2CA and CRP levels in the total sample (P=6.4×10^−4^), whereas PPP2CB showed a trend of association with the CRP level (**Table 4**). C4B level was correlated with Aβ_42_ level (P=0.047), whereas pTau231 level was significantly correlated with levels of C4A (P=0.023) and CRP (P=0.0024) (**Table S13**).

**Table 3.** Mean level of proteins measured by immunoassay in the FHS/BUADC dataset by each *APOE* genotype

| Outcome | ALL | | | $\epsilon 2/\epsilon 3$ | | | $\epsilon 3/\epsilon 3$ | | | $\epsilon 3/\epsilon 4$ | | | $\epsilon 4-$ vs $\epsilon 4+$ | | |
| --- | --- | --- | --- | --- | --- | --- | --- | --- | --- | --- | --- | --- | --- | --- | --- |
| | N | Mean | SD | N | Mean | SD | N | Mean | SD | N | Mean | SD | $\beta$ | SE | P-value |
| Aβ42 | 224 | 0.01 | 1.00 | 44 | -0.39 | 1.11 | 112 | 0.03 | 0.94 | 32 | 0.43 | 1.01 | 0.53 | 0.19 | 5.3E-03 |
| pTau181 | 165 | -0.04 | 0.97 | 32 | -0.32 | 0.97 | 84 | 0.03 | 0.97 | 24 | 0.08 | 0.97 | 0.20 | 0.21 | 3.6E-01 |
| pTau231 | 221 | 0.00 | 0.98 | 44 | -0.03 | 0.93 | 109 | 0.01 | 0.98 | 32 | 0.17 | 1.10 | 0.18 | 0.19 | 3.3E-01 |
| tTau | 221 | -0.01 | 0.99 | 44 | 0.25 | 1.21 | 109 | -0.15 | 0.96 | 32 | -0.14 | 0.85 | -0.07 | 0.19 | 7.1E-01 |
| pTau181/tTau ratio | 157 | 0.00 | 1.00 | 35 | -0.09 | 1.26 | 83 | 0.04 | 0.95 | 26 | -0.04 | 0.90 | 0.03 | 0.21 | 9.0E-01 |
| pTau231/tTau ratio | 219 | -0.01 | 0.98 | 44 | -0.20 | 0.96 | 107 | 0.06 | 0.93 | 32 | 0.27 | 0.96 | 0.34 | 0.18 | 5.9E-02 |
| PPP2CA | 203 | 0.00 | 1.00 | 38 | -0.14 | 1.19 | 104 | 0.01 | 0.97 | 31 | -0.01 | 0.97 | 0.00 | 0.20 | 9.9E-01 |
| PPP2CB | 203 | 0.00 | 1.00 | 38 | 0.07 | 0.96 | 104 | 0.06 | 1.06 | 31 | -0.45 | 0.92 | -0.52 | 0.19 | 8.0E-03 |
| C1q | 184 | -0.01 | 1.00 | 24 | 0.00 | 1.27 | 107 | 0.01 | 0.99 | 32 | -0.18 | 0.85 | -0.22 | 0.19 | 2.5E-01 |
| C4A | 203 | 0.00 | 1.00 | 38 | -0.16 | 0.99 | 104 | 0.01 | 0.96 | 31 | 0.17 | 1.08 | 0.18 | 0.19 | 3.3E-01 |
| C4B | 203 | 0.00 | 1.00 | 38 | 0.04 | 1.23 | 104 | 0.03 | 0.94 | 31 | -0.14 | 1.02 | -0.22 | 0.19 | 2.6E-01 |
| CRP | 214 | 0.02 | 0.98 | 41 | 0.00 | 0.82 | 110 | 0.16 | 0.92 | 32 | 0.18 | 0.99 | 0.02 | 0.17 | 9.2E-01 |

Immunoassays were conducted on prefrontal cortex area tissue. N: total sample size; SD: standard deviation

**Table 4.** Association of levels of PPP2CA and PPP2CB proteins with classical complement and AD-related proteins by *APOE* genotype

| Test: Outcome | ALL | | | $\epsilon 2/\epsilon 3$ | | | | $\epsilon 3/\epsilon 3$ | | | | $\epsilon 3/\epsilon 4$ | | | |
| --- | --- | --- | --- | --- | --- | --- | --- | --- | --- | --- | --- | --- | --- | --- | --- |
| | $\beta$ | SE | P-value | N | $\beta$ | SE | P-value | N | $\beta$ | SE | P-value | N | $\beta$ | SE | P-value |
| **PPP2CB:** |  |  |  |  |  |  |  |  |  |  |  |  |  |  |  |
| Aβ42 | -0.11 | 0.07 | 0.13 | 53 | -0.19 | 0.18 | 0.31 | 118 | 0.13 | 0.09 | 0.13 | 45 | -0.58 | 0.18 | **2.4x10<sup>-3</sup>** |
| pTau181 | -0.11 | 0.08 | 0.20 | 34 | -0.11 | 0.20 | 0.60 | 90 | -0.05 | 0.11 | 0.63 | 30 | -0.31 | 0.22 | 0.17 |
| pTau231 | -0.06 | 0.07 | 0.38 | 53 | -0.15 | 0.17 | 0.38 | 115 | -0.05 | 0.09 | 0.61 | 45 | -0.11 | 0.22 | 0.63 |
| tTau | 0.12 | 0.07 | 0.10 | 53 | 0.13 | 0.21 | 0.53 | 115 | 0.20 | 0.09 | **0.03** | 45 | -0.19 | 0.16 | 0.25 |
| pTau181/tTau | 0.13 | 0.08 | 0.10 | 30 | 0.46 | 0.23 | **0.05** | 82 | 0.12 | 0.10 | 0.25 | 26 | 0.05 | 0.20 | 0.81 |
| pTau231/tTau | -0.13 | 0.07 | 0.07 | 53 | -0.05 | 0.16 | 0.74 | 113 | -0.13 | 0.09 | 0.13 | 45 | -0.04 | 0.19 | 0.83 |
| PPP2CA | 0.35 | 0.07 | **2.4x10<sup>-7</sup>** | 53 | 0.66 | 0.17 | **4.9x10<sup>-4</sup>** | 118 | 0.31 | 0.09 | **4.4x10<sup>-4</sup>** | 45 | 0.32 | 0.18 | 0.09 |
| CRP | 0.12 | 0.07 | 0.09 | 48 | 0.37 | 0.15 | **0.02** | 113 | 0.14 | 0.09 | 0.10 | 42 | 0.22 | 0.19 | 0.27 |
| C1q | 0.10 | 0.07 | 0.16 | 30 | 0.35 | 0.26 | 0.20 | 107 | 0.05 | 0.10 | 0.60 | 39 | 0.18 | 0.17 | 0.28 |
| C4A | 0.11 | 0.07 | 0.12 | 53 | 0.14 | 0.17 | 0.42 | 118 | 0.10 | 0.09 | 0.25 | 45 | -0.03 | 0.22 | 0.89 |
| C4B | 0.35 | 0.07 | **3.3x10<sup>-7</sup>** | 53 | 0.49 | 0.19 | **0.01** | 118 | 0.28 | 0.08 | **8.7x10<sup>-4</sup>** | 45 | 0.59 | 0.17 | **1.8x10<sup>-3</sup>** |
| **PPP2CA:** |  |  |  |  |  |  |  |  |  |  |  |  |  |  |  |
| Aβ42 | -0.06 | 0.07 | 0.42 | 53 | 0.17 | 0.15 | 0.25 | 118 | -0.11 | 0.09 | 0.26 | 45 | -0.48 | 0.18 | **9.9x10<sup>-3</sup>** |
| pTau181 | -0.05 | 0.08 | 0.54 | 34 | -0.03 | 0.16 | 0.85 | 90 | -0.05 | 0.11 | 0.64 | 30 | 0.09 | 0.22 | 0.68 |
| pTau231 | 0.18 | 0.07 | **0.01** | 53 | -0.05 | 0.13 | 0.72 | 115 | 0.17 | 0.10 | 0.08 | 45 | 0.55 | 0.18 | **5.3x10<sup>-3</sup>** |
| tTau | 0.02 | 0.07 | 0.83 | 53 | 0.14 | 0.17 | 0.43 | 115 | 0.01 | 0.10 | 0.90 | 45 | -0.26 | 0.15 | 0.10 |
| pTau181/tTau | -0.04 | 0.08 | 0.59 | 30 | -0.07 | 0.19 | 0.72 | 82 | 0.00 | 0.10 | 0.99 | 26 | -0.15 | 0.19 | 0.43 |
| pTau231/tTau | 0.03 | 0.07 | 0.71 | 53 | -0.17 | 0.12 | 0.17 | 113 | 0.08 | 0.09 | 0.39 | 45 | 0.26 | 0.18 | 0.16 |
| CRP | 0.23 | 0.07 | **6.4x10<sup>-4</sup>** | 48 | 0.10 | 0.11 | 0.38 | 113 | 0.24 | 0.09 | **0.01** | 42 | 0.43 | 0.17 | **0.02** |
| C1q | -0.03 | 0.08 | 0.71 | 30 | -0.02 | 0.22 | 0.92 | 107 | -0.02 | 0.10 | 0.83 | 39 | 0.17 | 0.16 | 0.30 |
| C4A | 0.10 | 0.07 | 0.15 | 53 | -0.05 | 0.13 | 0.70 | 118 | 0.10 | 0.10 | 0.28 | 45 | 0.27 | 0.20 | 0.19 |
| C4B | 0.12 | 0.07 | 0.08 | 53 | 0.20 | 0.16 | 0.23 | 118 | 0.05 | 0.10 | 0.58 | 45 | 0.30 | 0.19 | 0.13 |

Protein levels were assessed in prefrontal cortex from FHS/BUADC subjects.
Nominally significant P-values (P<0.05) highlighted in bold.

### Confirmation of PPP2CB and C4A/B Correlation in Co-Culture System with Human Isogenic APOE iPSC-Derived Neurons and Astrocytes

We derived neurons (iNeurons) and astrocytes (iAstrocytes) using isogenic *APOE* KO, ε2/ε2 (APOE2), ε3/ε3 (APOE3) and ε4/ε4 (APOE4) human induced pluripotent stem cell (iPSC) lines (**Fig. S11a**) and we generated rapidly induced excitatory iNeurons within 4 weeks by overexpression of neuronal transcriptional factor neurogenin-2 (**Fig. 1a**, red). To generate iAstrocytes, we differentiated iPSCs into neural progenitor cells (NPCs) (**Fig. S11b**), and subsequently induced into iAstrocytes. Human iNeurons with isogenic *APOE* genotypes were co-cultured with the isogenic iAstrocytes for 9 days starting from Day 15 using cell inserts assembled in the 6-well plates (**Fig. 1b**).

**Fig. 1.**
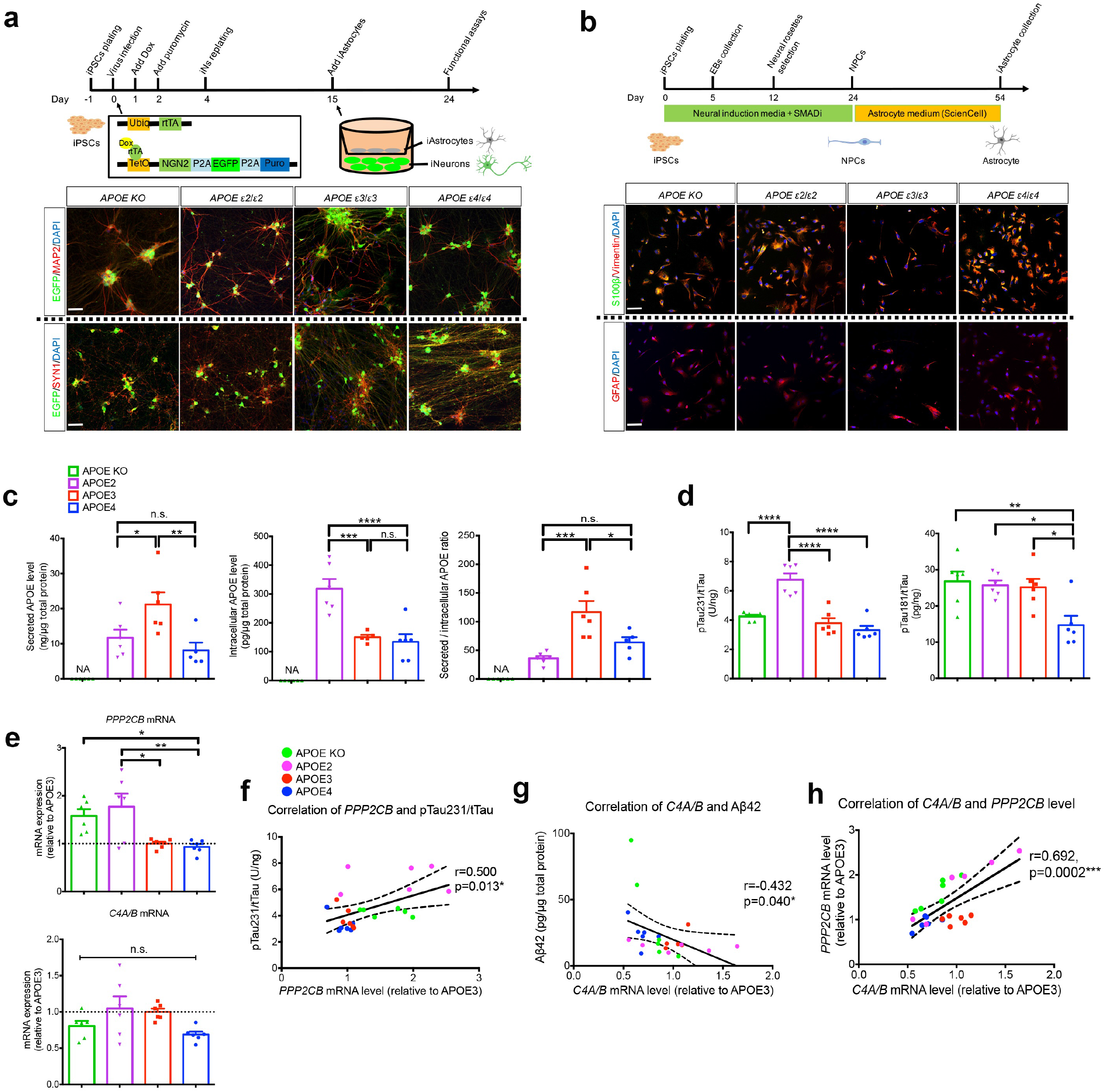
Association of PPP2CB and C4A/B gene expression with AD-related protein levels in isogenic *APOE* human iPSC-derived neurons co-cultured with astrocytes. **(a)** The scheme of generating iPSC-induced neurons and immunocytochemistry with MAP2 and synapsin1 (SYN1) antibodies in neurons. Scale bar, 75 μm. **(b)** The scheme of generating iPSC-derived astrocytes and immunocytochemistry with astrocytic markers (S100β, Vimentin, and GFAP). Scale bar, 75 μm. **(c)** Secreted APOE in conditioned media from iPSC-neurons/astrocytes co-cultures and intracellular APOE from iAstrocytes were measured by ELISA. n=5∼6 independent cultures per cell line. **(d)** Levels of pTau231/tTau and pTau181/tTau in iPSC-derived neurons were measured by quantitative ELISA. n=6 independent cultures per cell line. **(e)** Expression levels of *PPP2CB* and *C4A/B* genes by qRT-PCR and normalized to APOE3 neurons. n=6 independent cultures per cell line. Correlations of **(f)** *PPP2CB*expression with pT231/total Tau, **(g)** *C4A/B* expression with Aβ42, and **(h)** co-expression of *PPP2CB* with *C4A/B* in total iPSC-neurons samples. The dashed line indicates 95% confidence band of the best-fit line. n=24. Data expressed as mean ± s.e.m., one-way ANOVA with Tukey’s post hoc test, two-sided. Pearson correlation coefficients are shown. *p < 0.05, **p < 0.01, ***p < 0.001, ****p < 0.0001.

Since astrocytes mainly produce and secrete brain APOE to the extracellular space [31, 32], we measured the amount of intracellular and secreted APOE levels from APOEKO, APOE2, APOE3 or APOE4carrying iAstrocytes in the neuron-astrocyte co-culture system. APOE2 iAstrocytes exhibited the highest intracellular APOE level than other iAstrocytes, while we observed no APOE production in APOE KO iAstrocytes and significant reduction of APOE secretion in conditioned media from APOE2 and APOE4 compared to that from APOE3 iAstrocytes (**Fig. 1c**). The influence of *APOE* genotypes on AD-related proteins including tTau, pTau181, pTau231, Aβ40, and Aβ42 were quantified from APOEKO and different APOE carrying iNeurons. We observed the highest level of pTau231/tTau ratio from APOE2 while the pTau181/tTau ratio was the lowest from APOE4 iNeurons, compared that from APOE KO or other APOE carrying iNeurons (**Fig. 1d)**. There were no significant differences of Aβ (Aβ40 and Aβ42) levels among different *APOE* iNeurons (**Fig. S11c**).

Co-expression of *PPP2CB* with *C4A/B* was investigated in isogenic *APOE* iNeurons in the neuron-astrocyte co-culture system. Expression of *PPP2CB*was significantly increased in APOE2 compared to APOE3 and APOE4 iNeurons, while there were no significant differences in *C4A/B* expression among the APOE-defined groups of iNeurons (**Fig. 1e**). We confirmed the positive correlation of *PPP2CB*expression with pTau231/tTau ratio (P=0.013) but not with pTau181/tTau ratio (P=0.74) (**Fig. 1f and Fig. S11d**). *C4A/B* expression was not correlated with levels of pTau231 or pTau181 (**Fig. S11d**), while *C4A/B* expression was inversely correlated with Aβ42level (**Fig. 1g**). Taken together, we validated the findings in human brain tissue by confirming the correlation between *PPP2CB* and *C4A/B* expression (P=2.0×10^−4^) in iNeurons co-cultured with isogenic iAstrocytes regardless of the *APOE* genotype of the iNeuron (**Fig. 1h**).

## DISCUSSION

Results from this study provide evidence that the mechanisms underlying the deleterious effect of *APOE* ε4 and the protective effect of *APOE* ε2 on AD risk likely involve distinct biological pathways. Substantial evidence suggest that ε4 binds Aβ more aggressively than ε3 leading to greater deposition and less effective clearance of Aβ [33, 34]. A previous study showed that in the presence of Aβ ε4 promotes tangle pathology while ε2 is associated with fewer tangles, but both ε4 and ε2 are not associated with tangle pathology in the absence of Aβ [35]. Importantly, the role of *APOE* ε2 in AD has not been intensively studied and is often assumed to mirror the action of ε4 because it is associated with less Aβ [35]. Rather, our findings suggest that ε2 may regulate the interaction between the catalytic subunit of protein phosphatase 2A (PP2A) and complement component 4. Multiple independent lines of evidence obtained by an integrated systems biology approach support this idea. A near genome-wide significant association with a SNP in *PPP2CB* emerged as one of the top findings in an AD GWAS conducted among *APOE* ε2 carriers, and subsequent network analysis identified *PPP2CB* as a hub gene in ε2-related and biologically linked gene networks. Analysis of multiple omics data derived from brains donated by AD cases and controls including bulk and single nuclei RNA sequencing data, DNA methylation of CpG sites, and measures of AD-related proteins and PP2A catalytic subunit proteins yielded findings that further substantiate an *APOE* ε2-mediated role of PP2A subunit genes in AD.

In a separate study, we found that *C4A* and *C4B* were the most significantly differentially expressed genes in brain tissue from AD cases and controls with the *APOE* ε2/ε3 genotype. In the current study, we showed that levels of C4B and PPP2CB proteins were significantly correlated. To validate the *APOE* genotype-dependent effects of these proteins with respect to AD, we established a human iPSC-based co-culture system by deriving isogenic *APOE* iNeurons and iAstrocytes [30, 32]. We validated the correlated expression of*PPP2CB* with *C4A/B* as well as the correlation of *PPP2CB* expression with pTau231/tTau in iNeurons. Our results also showed that APOE2 isoform level was significantly associated with pTau231/tTau ratio, confirming a role of APOE2 in tau-related processes [36]. Our findings also suggest the possibility that PP2A dysfunction triggers activation of several components of the classical complement pathway leading to increased Tau phosphorylation and downstream sequela of tangle formation and neuronal death [36], whereas the APOE2 isoform may attenuate the effect of the classical complement cascade on events leading to AD [25].

Further scrutiny of the PP2A subunit behavior showed that brain levels of both PPP2CA and PPP2CB were negatively correlated with Aβ42 among ε4 carriers. These findings suggest that PP2A enzymatic activity under the elevated amyloid burden is likely disrupted leading to increased tau phosphorylation and that the two catalytic subunits are complementary at both mRNA and protein levels. Previously, it was shown that *PPP2CA*expression in the hippocampus [37] and PP2A activity in frontal and temporal areas [38] are significantly reduced in persons with AD. PP2A/Bα (brain PP2A enzyme) binds to tau at residues 221-396 with greater affinity for tau isoforms containing the adult 4 repeat (4R) than fetal tau [39]. *MAPT* (the gene encoding tau) missense mutations associated with frontotemporal dementia with parkinsonism-17 (FTDP-17) reside in the microtubule-binding domain and inhibit PP2A binding by 20-70% [40]. PP2A enzymatic activity is negatively correlated with tau phosphorylation levels at multiple sites in human brain and this association is greater at Tau position 231 than 181 [41]. Phosphorylation at the T231 site of Tau protein significantly decreases binding of tau to PP2A leading to poor dephosphorylation activity by the brain PP2A enzyme [42, 43]. These reports are consistent with our finding of significant correlation of the amount of pTau231 but not pTau181 with of PP2A catalytic subunit protein levels (PPP2CA and PPP2CB) in human brains, as well as with observed expression patterns of *PPP2CB* in isogenic APOE human iNeurons and iAstrocytes. A previous study found that cerebrospinal fluid (CSF) pTau231 compared to pTau181 level better differentiates controls from AD patients with higher specificity (86% vs. 46%) at identical sensitivity (85%) [44]. The investigators of that study also observed that prediction in the diagnostic classification with pTau231 but not pTau181 is sensitive with *APOE* ε4 carrier status [44]. We found that *APOE* ε4 carrier status was significantly associated with Aβ_42_ and nearly so with pTau231/tTau ratio, but not associated with pTau181/tTau ratio.

Other regulators of brain PP2A include leucine carboxyl methyltransferase 1 (LCMT-1) which activates PP2A by methylation of the catalytic C subunit at the L309 site and protein phosphatase methylesterase 1 (PME-1) which inhibits PP2A by demethylating the L309 site [45, 46]. Expression of the PP2A catalytic subunit L309A mutation increases tau phosphorylation in transgenic mice [47]. Proteomic analysis of exosomes from human induced pluripotent stem cell (iPSC)-derived neurons expressing mutant Tau (P301L or V337M) identified ANP32A, an endogenous PP2A inhibitor, present exclusively in tau-mutant exosomes [48]. Potential clinical implications of our finding on inverse relationships between levels of catalytic subunits of PP2A and the Aβ42 level is indirectly supported by the observation that the heterozygous *PME-1* knockout mice which display up-regulated PP2A activity are resistant to Aβ-induced cognitive impairment [49]. Direct activation of PP2A in rat hippocampus by a tricyclic sulfonamide treatment increased PP2A activity, decreased tau phosphorylation and Aβ42/Aβ40 levels in cells, and prevented neuronal synapse impairment [50]. Taken together, our study of human brain tissue and recent AD cell and mouse model studies support a therapeutic concept for developing or repurposing PME-1 inhibitors and PP2A activators [51] to slow progression of neurodegeneration and improve cognitive function.

Complement 4 is a key component of the classical complement pathway, an innate immune system. *C4A* and *C4B* are among the group of major histocompatibility complex (MHC) III genes that is located between MHC I and MHC II gene clusters. The MHC locus has been consistently reported as one of the most significant association signals for late onset AD [9, 52]. However, it has been challenging to disentangle variants in this region that are in high linkage disequilibrium (LD). A previous study showed a statistically significant increase in the repeat length of copy number variants in both *C4A* and *C4B* in AD compared to control subjects [53]. Nerl et al reported a high relative risk (RR=8.8) of AD associated with a *C4B* variant in a study of 42 AD cases and 59 age-matched controls [54], but other studies were unable to confirm this finding [55, 56]. This observation, together with evidence showing reduced synaptic pruning in C4 knockout mice [57], suggests that an increased amount of C4 protein in the synapse promotes pruning.

Multiple lines of evidence from mouse models link ApoE to classical complement proteins. It has been shown that ApoE isoforms bind directly to C1q protein and modulate complement-dependent synapse loss, inflammation, microglia accumulation, and atherosclerosis [25]. In another study, the rate of synapse pruning by astrocytes varied according to their enrichment of a specific ApoE isoform (E2>E3>E4). In addition, C1q protein accumulation in the hippocampus from aged human-*APOE*2/3/4 knock-in mice was isoform dependent (E2<E3<E4) [58]. Although there is no direct evidence linking PP2A to the classical complement cascade, especially in an *APOE* genotype dependent manner, perturbed interactions of ApoE, PP2A and complement proteins C4A/B and C1qmay promote AD-related pathological changes in brain.

In addition to *PPP2CB*, among the top-ranked *APOE* ε2 network genes that were linked to AD by findings from analyses of brain omics data, expression of *STAT3* (signal transducer and activator of transcription 3) only was significantly associated with pTau231/tTau ratio (P=2.7×10^−4^). *STAT3* is activated in reactive astrocytes in transgenic mouse models of AD [59], and inhibition of its expression prevents astrogliogenesis and promotes neurogenesis in a brain inflammation-induced human neural progenitor cell model [60]. A gene co-expression network analysis of data derived from peripheral blood obtained by AD, mild cognitive impairment (MCI), and control subjects revealed that *STAT3* is a hub gene for ubiquitin degradation, inflammatory signaling, and insulin resistance pathways and suggested that its expression may mediate the pathological changes associated with MCI [61].

Among other top-ranked genes that emerged from the *APOE* ε2-related GWAS, we identified a genome-wide significant interaction between *CGNL1* SNP rs17239735 and ε2. The connection of *CGNL1* to AD was supported by findings from analyses of multiple brain omics data. *CGNL1* encodes a member of the cingulin family, which localizes to adherens and tight cell-cell junction and regulates the activity of the small GTPases RhoA and Rac1 (http://genecards.org). In mice, the Cgnl1 protein is enriched in CNS endothelial cells in the blood-brain barrier [62] and perturbed during traumatic brain injury [63]. We identified a genome-wide significant association of the interaction between *CGNL1* SNP rs17239735 and ε2. The effect of rs17239735 on AD risk among ε2 carriers was in the opposite direction among persons with ε3/ε3 or ε3/ε4 genotypes. We also observed that *CGNL1* expression was significantly greater in AD compared to control brains, and was significantly associated with amyloid plaque density.

Our findings should be considered in light of several caveats. First, the sample size of ε2 carriers especially among AD cases is small and thus reduced power for a GWAS in this *APOE* genotype group. This problem also hampered opportunities for replication. To substantiate our findings and evaluate statistical differences of effect direction across *APOE* genotype subgroups, we performed a GWAS including a term for the interaction between ε2 and SNP. Among the genes that were included in subsequent network analyses, comparisons of expression and methylation with AD and related measures, and proteomic studies, all but one gene showed significant association with AD (P<10^−5^) among ε2 carriers and for interaction with ε2. Second, differential expression analysis using single nuclei RNA sequencing data was limited by small numbers of expressed cells especially for microglia, endothelial cells, and pericytes. Except for results within neurons, the majority of highly relevant genes were not expressed or significant after FDR correction in other cell types. Therefore, our single nucleus expression findings are incomplete. Third, the small number of brains from *APOE* ε2 carriers in the FHS/BUADC sample limited our ability to evaluate fully *APOE*-genotype specific associations with AD-related proteins. Fourth, our finding of significant association of a *PPP2CB* variant with AD risk among ε2 carriers needs to be interpreted in the context of association of *PPP2CB* expression with the pTau231/tTau ratio among ε4 carriers and significant correlation of PPP2CB protein with Aβ42 levels. One possible explanation is that the ApoE4 isoform or elevated Aβ42 levels among ε4 carriers may exacerbate dysregulated *PPP2CB* expression leading to increased pTau231/tTau ratio, while the ApoE2 isoform may mitigate (inhibit) this consequence. The absence of association of *PPP2CB* with AD in ε4 carriers would be consistent because ApoE4 may not inhibit the effect of PPP2CB on phosphorylation of tau regardless of *PPP2CB* genotype, whereas ApoE2’s impact on tau can be modified by *PPP2CB* genotype. This hypothesis warrants further study. Fifth, our focus on mechanisms underlying the protective effect of ε2 obscured potentially important findings from other *APOE* genotype groups. In particular, because one of the ε3/ε4 networks featured complement pathway genes, C1q, C5, and CR1, complement activation may not be ε2-specific. Follow-up experiments of genes in the other networks are needed. Sixth, we aware of a recent study showing that pTau217 measured in plasma accurately discriminated AD from other neurodegenerative disorders [64], but were unable to assess this metabolite in our sample due to lack of a reliable assay tool. Finally, our co-culture system may not represent a true microenvironment of neurons and astrocytes in postmortem brains due to a lack of microglia. In fact, we failed to confirm APOE genotype dependent effects of amyloid-β in the co-culture system. In addition, we tested one isogenic line for iPSC experiments, since this was the only isogenic line available with the required *APOE* KO, ε2/ε2, ε3/ε3, and ε4/ε4 backgrounds for the experiments. Nonetheless, our newly developed co-culture system using isogenic *APOE* genotype-specific human iPSC-derived neurons and astrocytes demonstrated that this system is robust for assessing effects of *APOE* and *APOE* genotype-dependent changes related to AD.

## CONCLUSIONS

Currently the Food and Drug Administration have approved only four drugs for AD, the latest a decade ago, and to date no drugs effectively retard progression of the disease [65-67]. More than 400 AD clinical trials conducted between 2002 and 2012 failed, and no drugs were approved after 2003 [65-67]. Efforts in ApoE-targeted therapeutics including modification of the ApoE4 structure, modulation of ApoE lipidation, inhibiting ApoE and Aβ interaction, and development of ApoE-mimetic peptides have been largely unsuccessful [68]. Given the well-documented high correlation between tau and cognitive impairment in humans [8], our study suggests that modifying the interaction between PP2A and complement pathway components may reduce tau phosphorylation. Thus, extensive follow-up studies validating this novel link are warranted and may lead to novel drug development or repurposing existing PME-1 inhibitors, PP2A activators, or complement-targeted drugs for treating AD [51, 69].

## Data Availability

Genotypes and clinical/neuropathological phenotype data are accessible by directly applying to the collaboration agreement (ACT) or by their websites: ROSMAP at https://radc.rush.edu or www.synapse.org, and ADNI at http://adni.loni.usc.edu. Genotypes and clinical/neuropathological phenotype data from other studies can be accessed by applying directly to the National Institute on Aging Genetics of Alzheimer Disease Data Storage Site (NIAGADS), a NIA/NIH-sanctioned qualified-access data repository, under accession NG00075. Data supporting the findings of this study are available from the NIAGADS website (https://www.niagads.org/). The custom codes in R that generated the findings of this study are available by authors upon request.

Http://www.synapse.org

http://adni.loni.usc.edu

https://www.niagads.org

## DECLARATIONS

### Ethics approval and consent to participate

This study was approved by the Boston University Institutional Review Board (H-36875). Written informed consent was obtained from all participants. This study was performed in accordance with the Helsinki declaration.

### Consent for publication

Not applicable

### Availability of data and materials

Genotypes and clinical/neuropathological phenotype data are accessible by directly applying to the collaboration agreement (ACT) or by their websites: ROSMAP at https://radc.rush.edu or www.synapse.org, and ADNI at http://adni.loni.usc.edu. Genotypes and clinical/neuropathological phenotype data from other studies can be accessed by applying directly to the National Institute on Aging Genetics of Alzheimer’s Disease Data Storage Site (NIAGADS), a NIA/NIH-sanctioned qualified-access data repository, under accession NG00075. Data supporting the findings of this study are available from the NIAGADS website (https://www.niagads.org/). The custom codes in R that generated the findings of this study are available by authors upon request.

## Authors’ contributions

J.C., R.P., and J.H. performed genetic and brain omics data analyses. Y.Y. and T.I. designed and conducted human iPSC experiments. G.M., W.X., and T.D.S. conducted immunoassay experiments using FHS/BUADC autopsied brains. K.L.L. provided statistical advice. G.R.J., Y.Y., W.X., D.A.B., T.M.F., L-S. W., J.L.H., R.M., M.A.P-V. G.D.S., R.A., K.L.L., T.I., T.D.S., and L.A.F. reviewed and edited the manuscript. D.A.B., R.A., T.D.S. provided neuropathological data. G.R.J. and L.A.F. supervised the project. G.R.J., J.L.H., R.M., M.A.P-V., G.D.S., and L.A.F. obtained funding for the project.

## Acknowledgements

We thank contributors who collected samples used in this study, as well as patients and their families, whose help and participation made this work possible. We thank Dr. Marilyn Miller from NIA who is an *ex-officio* ADGC member.

