## Supplemental Materials for "Protein phosphatase 2A, complement component 4, and *APOE* genotype linked to Alzheimer’s disease using a systems biology approach"

**Supplementary Information**

**Supplementary Note**

**Bulk RNA Sequencing and Data Processing:**

Total RNAs from the dorsolateral prefrontal cortex (Brodmann area 8/9) of 208 brains from the FHS/BUADC study were extracted using the Promega Maxwell RSC simplyRNA Tissue Kit (Cat No# AS1340) according to the manufacturer's protocol. The integrity and quality of RNA (RNA integrity number, RIN) was determined using the Agilent 2100 Bioanalyzer with RNA 600 Nano Chips (Cat No# 5067-1511). After excluding brain samples with RIN < 5, brain samples were randomized into seven library batches based on diagnosis, *APOE* genotype, sex, and RIN. Since there were only seven samples from AD cases with *APOE* genotypes 2/2 or 2/3, these specimens were included in batches 1 to 3 only. The BU Microarray & Sequencing Resource Core performed RNA sequencing (RNA-seq) library preparation. The libraries were prepared from total RNA enriched for mRNA using NEBNext Poly(A) mRNA Magnetic Isolation Module and NEBNext Ultra II Directional RNA Library Preparation Kit for Illumina (New England Biolabs, USA) and sequenced on an Illumina NextSeq 500 instrument (Illumina, USA). A total of 193 of the 208 samples remained for mapping after pre-processing. The average coverage of the remaining samples was 50 million reads for the entire sample.

RNA-seq data from 193 FHS/BUADC brains were processed by our automated pipeline. We conducted quality control of the RNA-seq data for sequencing quality, over-abundance of adaptors, and over-represented sequence using the FastQC. Low-quality reads (5% of the total) were filtered out using the *Trimmomatic* option, which is a fast, multithreaded command line tool to trim and crop Illumina (FASTQ) data and remove adapters [1]. After trimming adapter sequences from read, we aligned the initial QC passed reads to human reference genome (GRCh38.95) using the STAR, which implements 2-pass mapping to increase mapping chances of splice reads from novel junctions [2, 3]. We used the *readFilesCommand* option for reading input files, the *TranscriptomeSAM* option for mapped reads translated into transcript coordinates, and the *GeneCounts* option for counting mapped reads per gene under the mapping mode set to *quantMode* and then *twoPassMode* options. This process produced a BAM file of mapped paired end reads for each sample with a corresponding alignment report file.

Post alignment quality of bam files was evaluated for gene coverage and junction saturation using the RSeQC [4]. The RSeQC program can comprehensively evaluate quality of mapped reads from RNA-seq data for each sample by evaluating uniformity of coverage over gene bodies using the *gene_Body_coverage* option, checking if inner distance between read pairs is within expecting range of fragments lengths using the *inner_distance* option, and ensuring sequencing depth using the *junction_saturation* option. Gene and isoform levels were quantified using the RSEM and Bowtie2 and annotated using the Homo_sapiens.GRCh38.95.gtf annotation files. This process generated gene or isoform expression data for each sample containing gene id, gene length, effective gene length, expected count, counts per million (CPM), and FPKM reads. We investigated batch effects of seven different library and three sequencing batches with the quantified gene expression data using principle component analysis (PCA). We did not detect significant batch effects for both library and sequencing batches. Gene expression levels were quantified as normalized fragments per kilobase of transcript per million (FPKM) reads, and genes with FPKM reads < 5 were excluded.

**Single Nuclei RNA Sequencing Data Processing and Differential Expression Data Analysis**

Droplet-based snRNA-seq libraries were constructed by Chromium Single Cell 3′ Reagent Kits v2 according to the manufacturer’s protocol and RNA sequencing was performed using a NextSeq 500/550 instrument and High Output v2 kits with a setting of 150 cycles [5]. Read counts were aligned to a reference genome (GRCh38) using the CellRanger software [6]. We used a 200 cut-off value for unique molecular identifiers (UMI) for better detection of small proportion of cell types in snRNA-seq data [5]. After excluding genes that are non-protein coding or with low expression across all cell types, the dataset used for downstream analyses contained 69,918 nuclei and 5,578 marker genes. Data were normalized and clustered using the Seurat program [7]. Normalized gene expression measures from each cell were scaled by 10,000 multiplied by the total library size and then log transformed using the ScaleData function in R. We conducted principle component analysis of cell type specific expression levels using data for previously established marker genes for each brain cell type [5] and t-distributed Stochastic Neighbor Embedding (tSNE) method [8]. The proportions of individual cells for endothelial (0.14%), pericytes (0.2%), microglia (2%), oligodendrocyte progenitor cells (OPC; 4%), and astrocytes (5%) were much smaller than those for excitatory neurons (50%), inhibitory neurons (12%), and oligodendrocytes (26%). Endothelial cells and pericytes were excluded from subsequent analyses due to their very low proportion.

**Fig. S1.** Study design

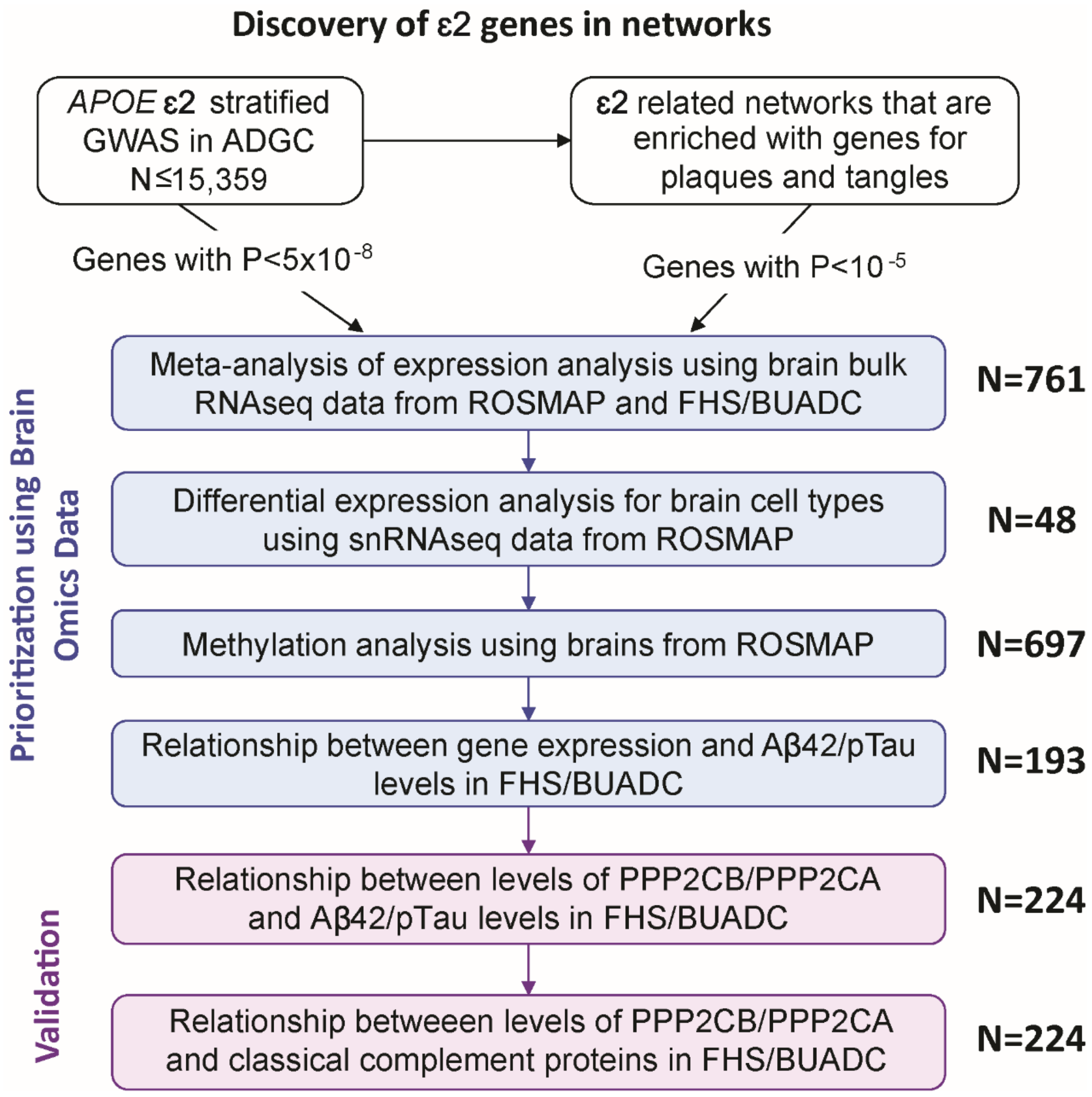

**Fig. S2.** Quantile-quantile plots for AD GWAS in *APOE* genotype subgroups. Observed (y-axis) vs. expected (x-axis) *P*-values for AD were plotted with association results using ε2 carriers **(a)**, ε2 interaction **(b)**, ε3/ε3 subjects **(c)**, ε3/ε4 subjects **(d)**, ε4/ε4 subjects **(e)**, and ε4 interaction **(f)**.

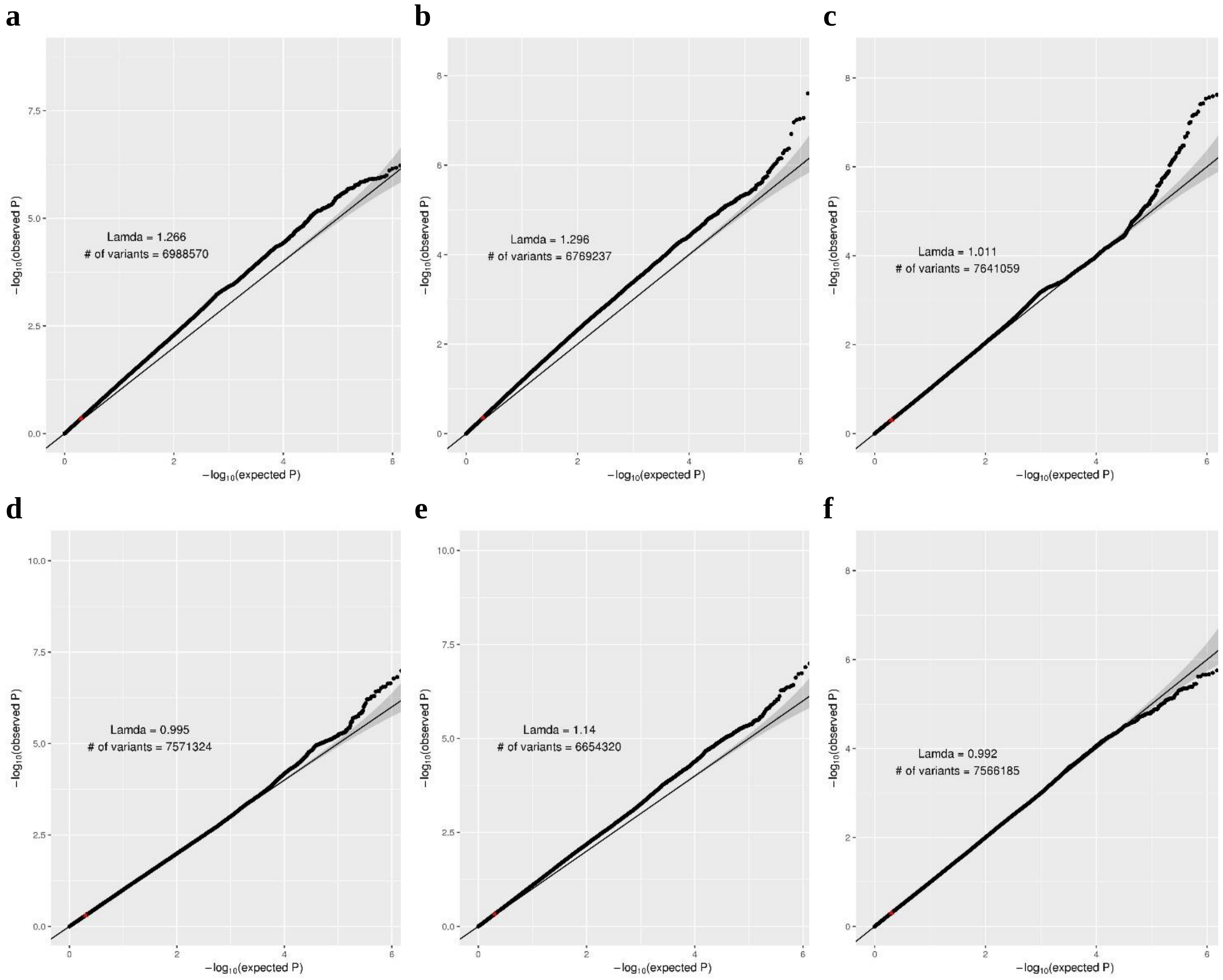

**Fig. S3.** Manhattan plots for AD GWAS in *APOE* genotype subgroups. Observed –log *P*-values (y-axis) in SNP-based tests for AD were plotted by chromosome (x-axis) using ε2 carriers **(a)**, ε2 interaction **(b)**, ε3/ε3 subjects **(c)**, ε3/ε4 subjects **(d)**, ε4/ε4 subjects **(e)**, and ε4 interaction **(f)**.

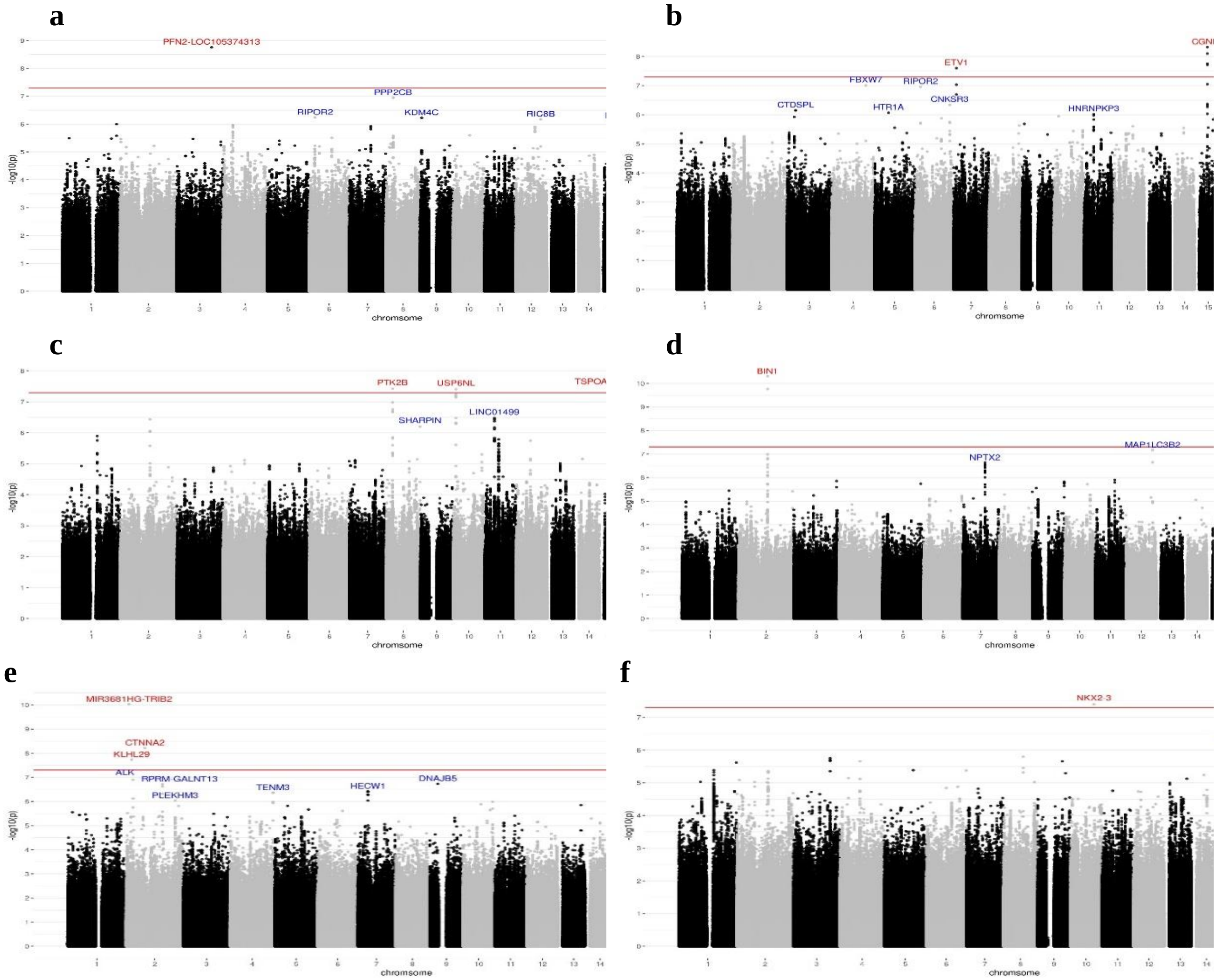

**Fig. S4**. Regional plots of loci showing significant evidence for association with AD risk ($P<10^{-7}$) in ε2 carriers and ε2 interaction tests. The observed –log *P*-value (y-axis) for each SNPs was plotted against chromosomal location in base pairs (x-axis). Computed estimates of linkage disequilibrium ($r^2$) of SNPs with the most significant SNP were derived from subjects of European ancestry. **(a)** *LOC105371413* region in ε2 carriers, **(b)** *PPP2CB* region in ε2 carriers, **(c)** *ETV1* region in ε2 interaction tests, and **(d)** *CGNL1* region in ε2 interaction tests.

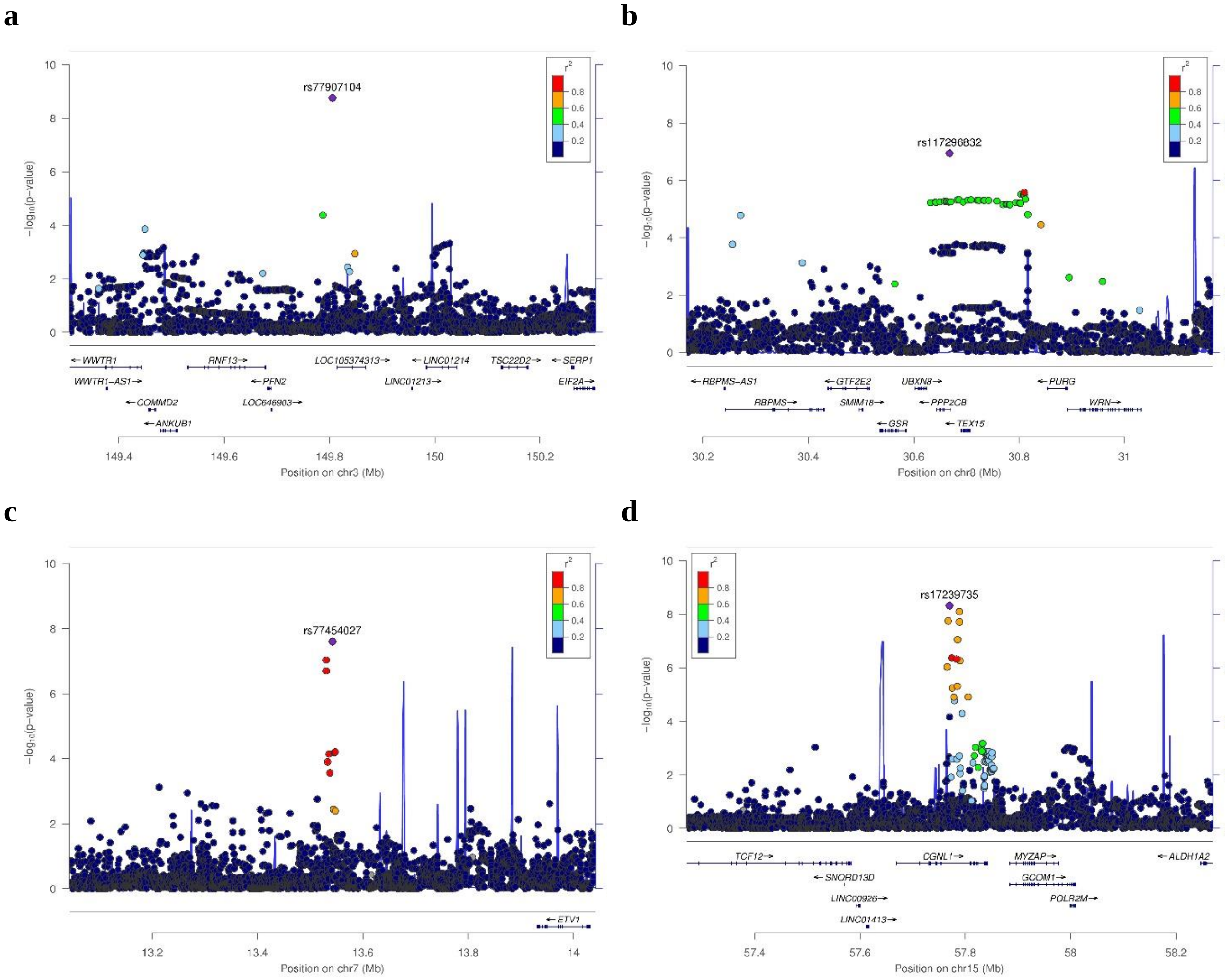

**Fig. S5**. Regional plot showing the association of *TRIM37* with AD risk among ε3/ε3 subjects. The observed –log *P*-value (y-axis) for each SNP was plotted against chromosomal location in base pairs (x-axis). Computed estimates of linkage disequilibrium ($r^2$) of SNPs with the most significant SNP were derived from subjects of European ancestry.

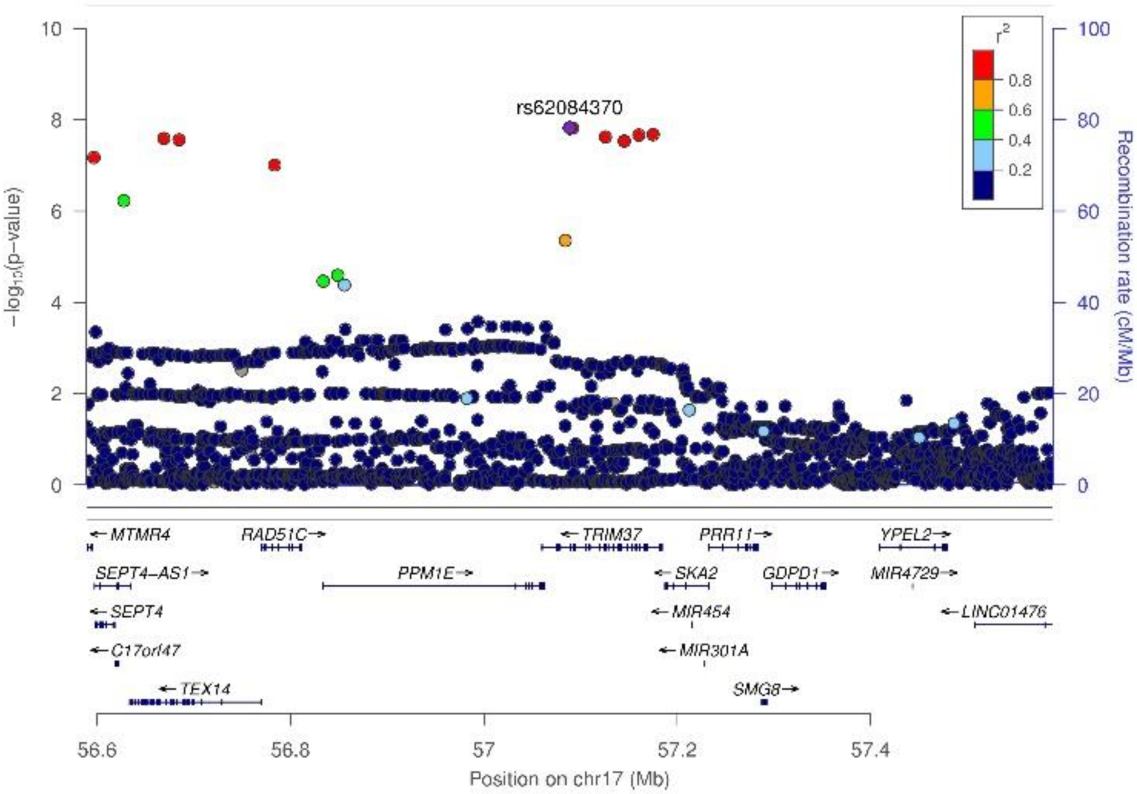

**Fig. S6**. Regional association plots for loci significantly associated with AD risk ($P<10^{-6}$) among ε4/ε4 subjects including **(a)** *MIR3681HG-TRIB2*, **(b)** *KLHL29*, **(c)** *CTNNA2*, and **(d)** *ZNF443*. The observed –log *P*-value (y-axis) for each SNP was plotted against chromosomal location in base pairs (x-axis). Computed estimates of linkage disequilibrium ($r^2$) of SNPs with the most significant SNP were derived from subjects of European Ancestry.

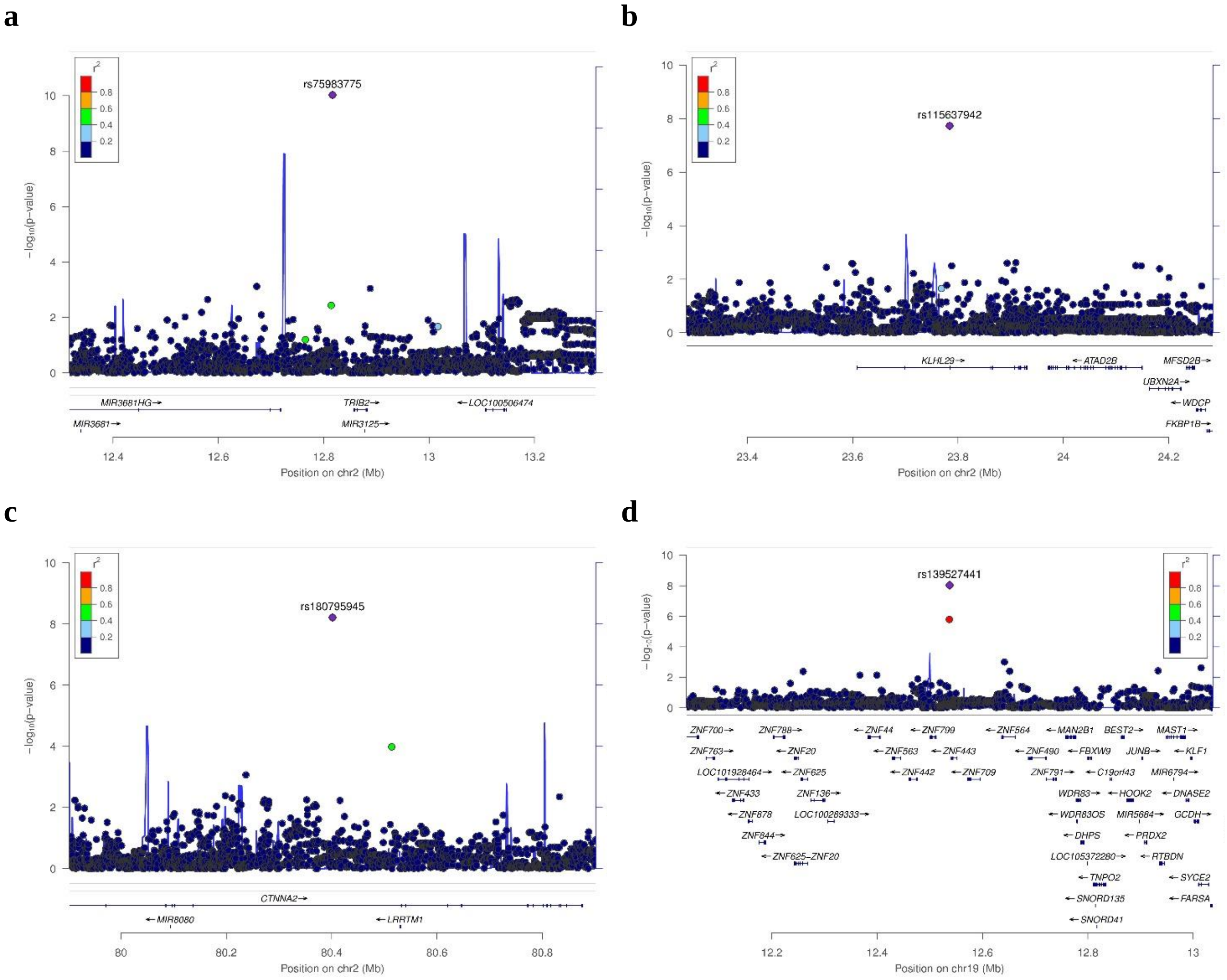

**Fig. S7**. Regional plot showing association of *NKX2-3* with AD risk in ε4-interaction tests. The observed –log *P*-value (y-axis) for each SNP was plotted against chromosome base pairs (x-axis). Computed estimates of linkage disequilibrium ($r^2$) of SNPs with the most significant SNP were derived from subjects of European ancestry.

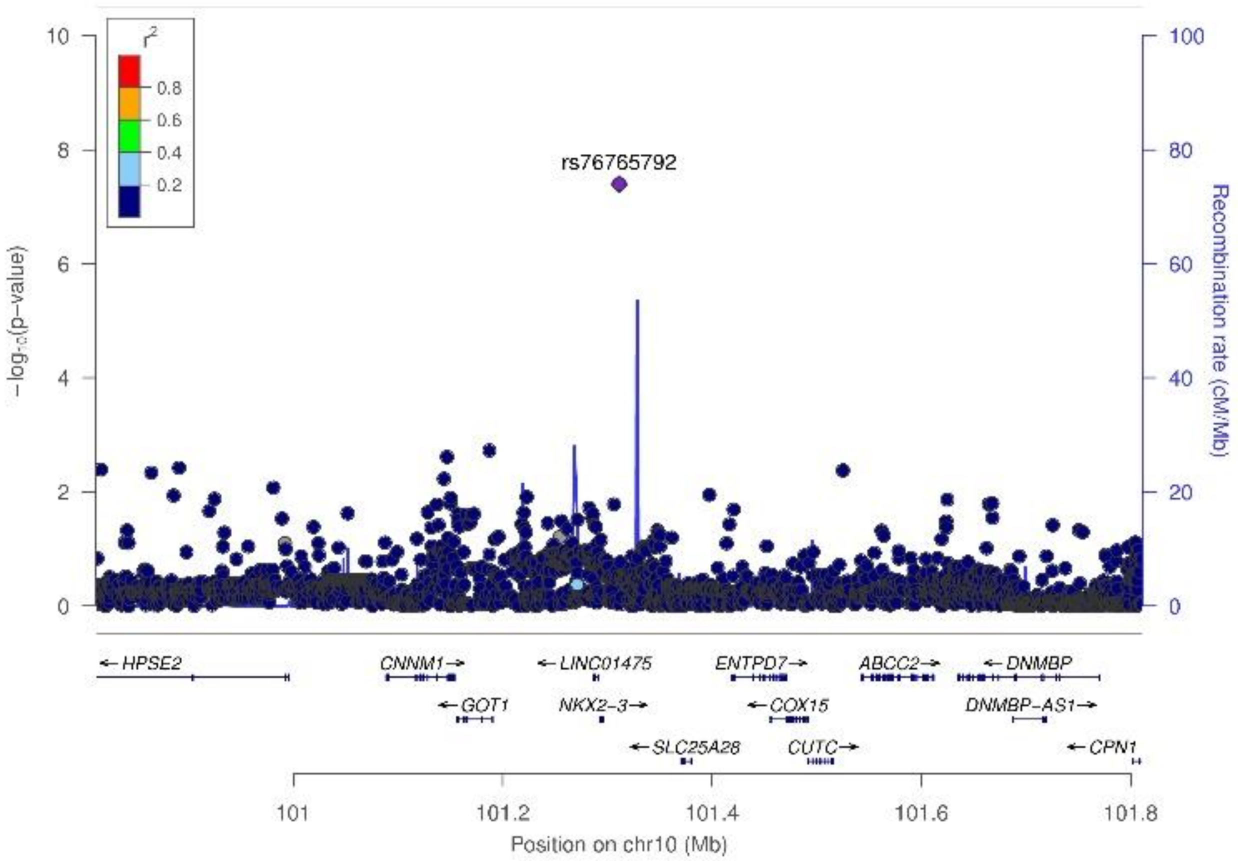

**Fig. S8.** *APOE* ε2 network enriched with genes associated with density of neuritic plaques and neurofibrillary tangles ($P<10^{-5}$). Gradient red color of the seed genes represents strength of P values for association with AD among *APOE* ε2 carriers. *PPP2CB* is a hub gene that is also the most significantly AD-associated gene in this network ($P=1x10^{-7}$).

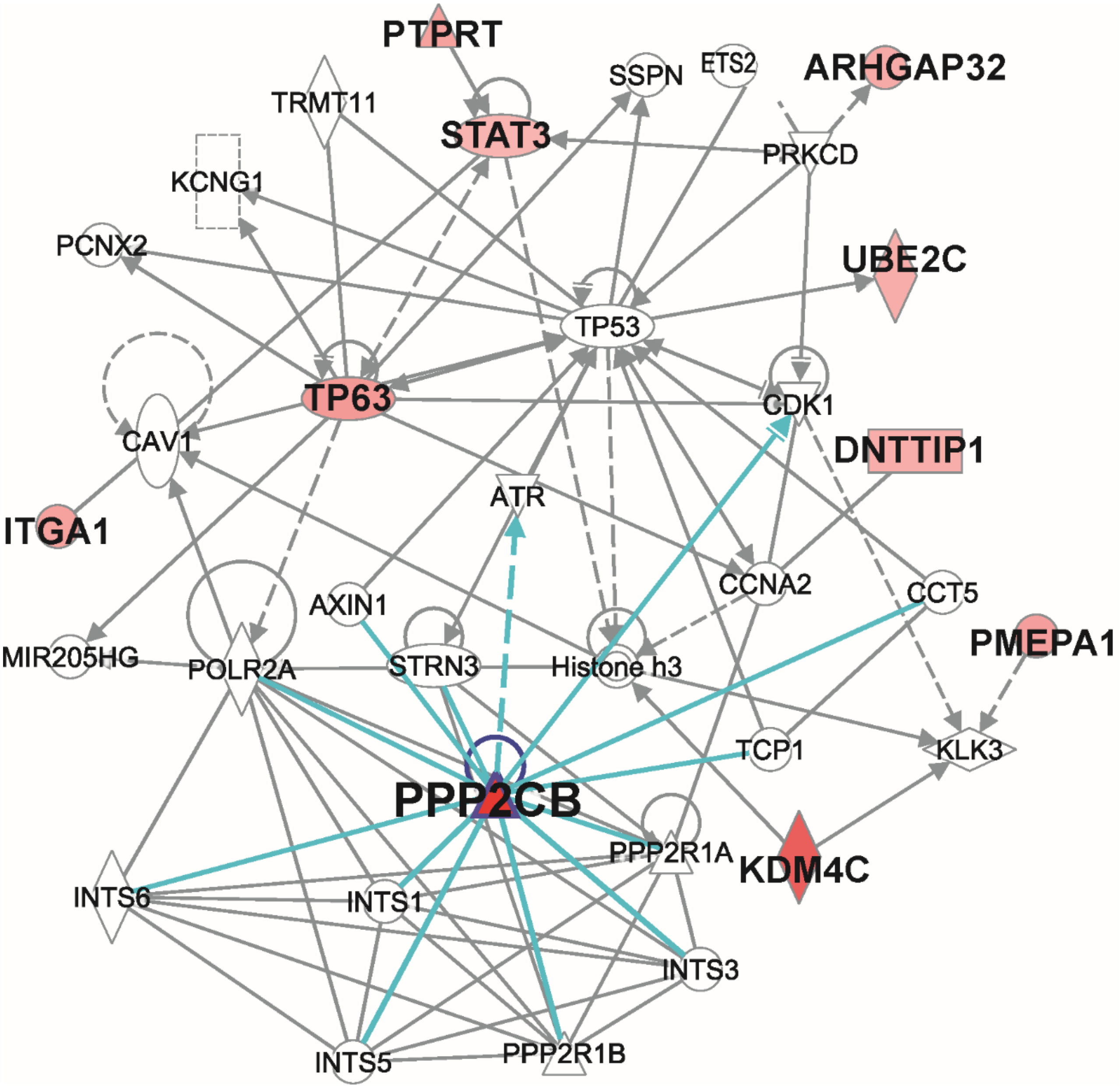

**Fig. S9.** *PPP2CB* expression profile in brain determined by single nuclei RNA sequencing (snRNA-seq). (**a**) Proportion of cells expressing *PPP2CB* in each cell type that represent at least 1% of all single cells examined; (**b**) Proportion of excitatory (ExN) and inhibitory (InN) neurons in each sample from AD cases and controls expressing *PPP2CB*; (**c**) Violin plots for distribution of *PPP2CB* expression levels in AD cases and controls in each type; (**d**) Two-dimensional tSNE projection of *PPP2CB* expression in the total sample of annotated cells. Ast: astrocyte; ExN: excitatory neuron; InN: inhibitory neuron; Mic: microglia; Oli: oligodendrocyte; OPC: oligodendrocyte progenitor cell.

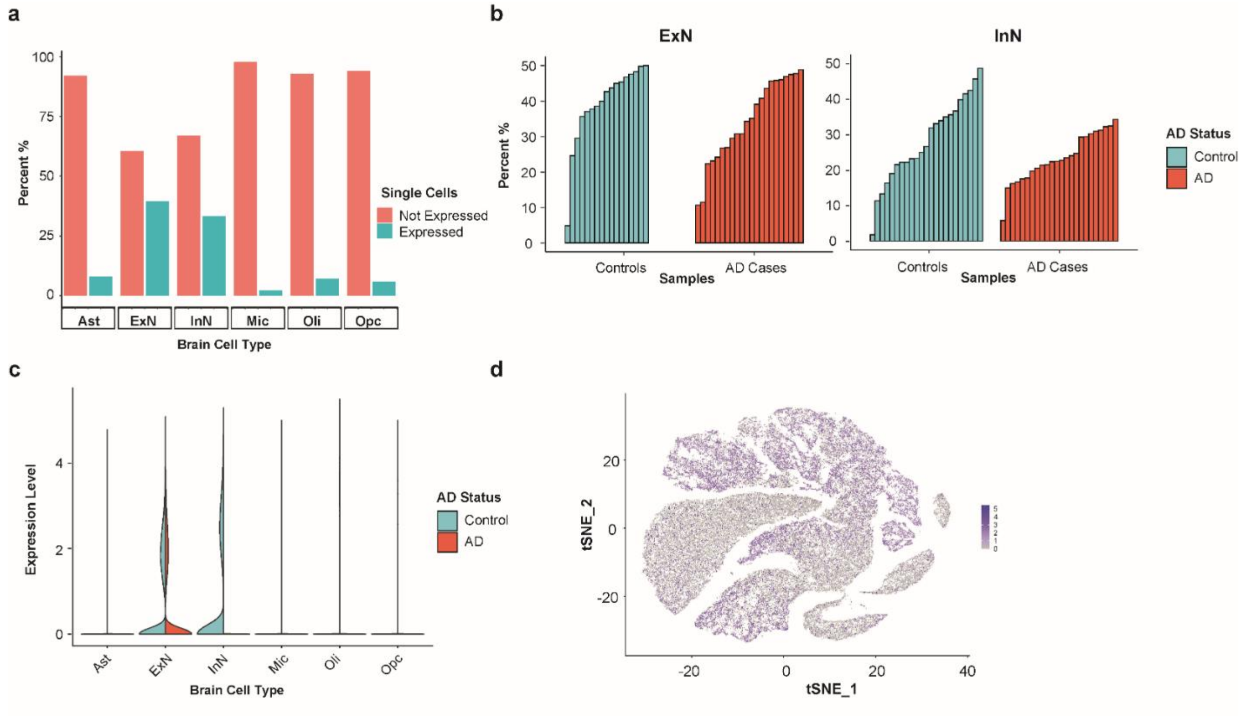

**Fig. S10.** Association of PPP2CB protein level with AD-related genes and proteins measured in the prefrontal cortex region of brains from FHS/BUADC subjects. Levels of PPP2CB, PPP2CA, amyloid beta-42 (Aβ42), and C4B were rank transformed after adjusting for age at death and sex. **(a)** Box plot showing the distribution of PPP2CB level in *APOE* ε4-negative (E4N) and ε4-positive (E4P) subgroups. Distributions were compared using linear regression. **(b-d)** PPP2CB level plotted against PP2A which is the PPP2CB enzyme complex **(b)**, $A\beta_{42}$ **(c)**, and C4B **(d)**. Fitted regression lines are shown for the total sample (ALL; black), *APOE* ε4-negative subjects (E4N; red), and ε4-positive subjects including ε3/ε4 and ε4/ε4 carriers (E4P; green). P-values indicate the significance level of the correlation of PPP2CB level with each trait in each group.

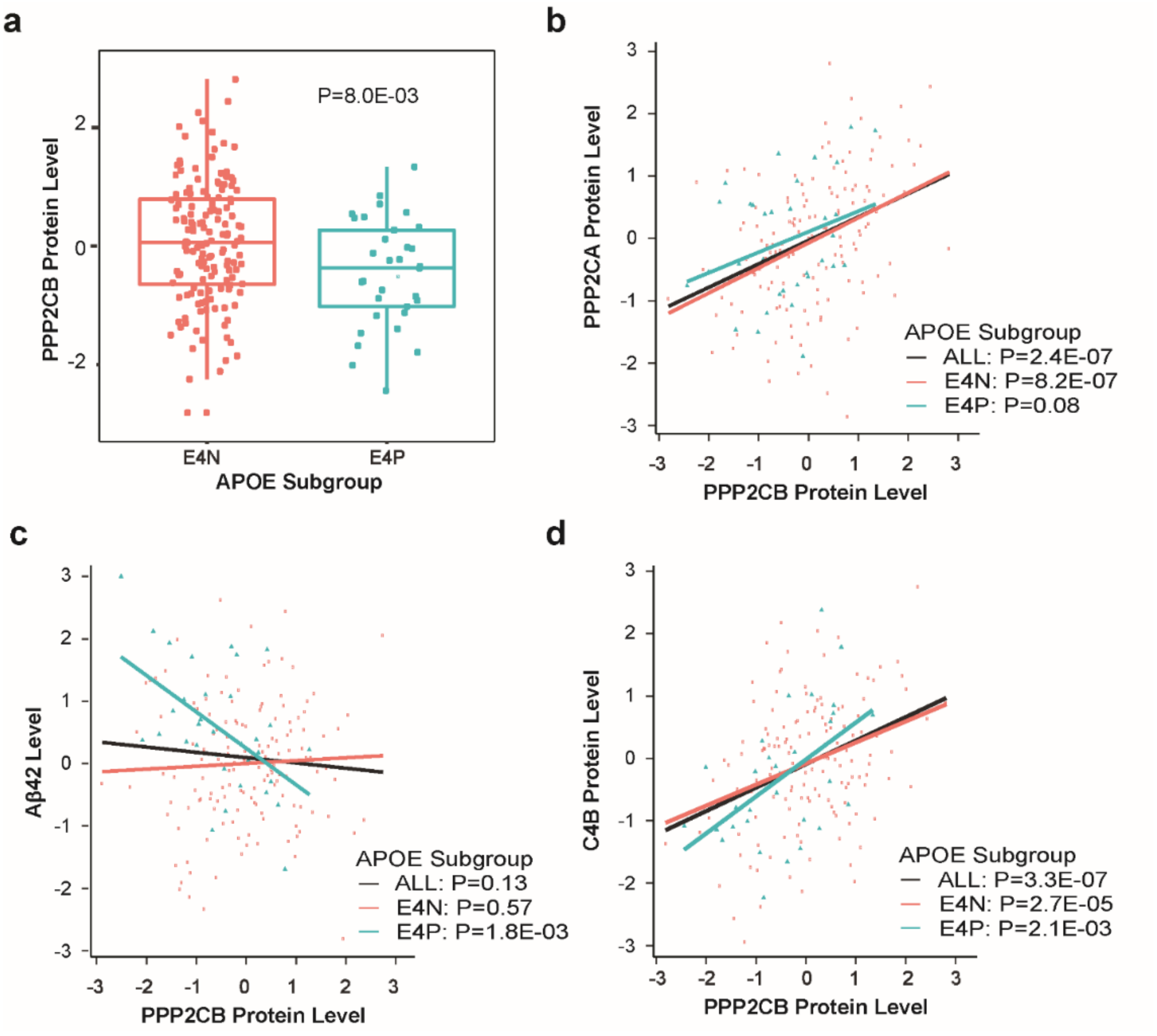

**Fig. S11.** Association of *PPP2CB* and *C4A/B* expression with AD-related protein levels in isogenic *APOE* human iPSC-derived neurons co-cultured with astrocytes. **(a)** Immunocytochemistry with pluripotent stem cell markers (OCT3/4, NANOG, Alkaline Phosphatase (AP) and SSEA-4). Scale bar, 75 μm. **(b)** Immunocytochemistry with neural progenitor cell markers (SOX2, NESTIN and PAX6). Scale bar, 75 μm. **(c)** Levels of total Tau, Aβ40, Aβ42 and Aβ42/Aβ40 in iPSC-derived neurons were measured by quantitative ELISA. n=5~6 independent cultures per cell line. **(d)** Correlation of *PPP2CB* expression with pT181/total Tau, *C4A/B* expression with pT231/total Tau, and *C4A/B* expression with pT181/total Tau in all iPSC-neurons. Dashed line indicates 95% confidence band of the best-fit line. n=24. Data expressed as mean ± s.e.m., one-way ANOVA with Tukey's post hoc test for total Tau and Aβ42/Aβ40, Kruskal-Wallis test with Dunn's post hoc test for Aβ40 and Aβ42, two-sided. r: Pearson correlation coefficient. *p < 0.05.

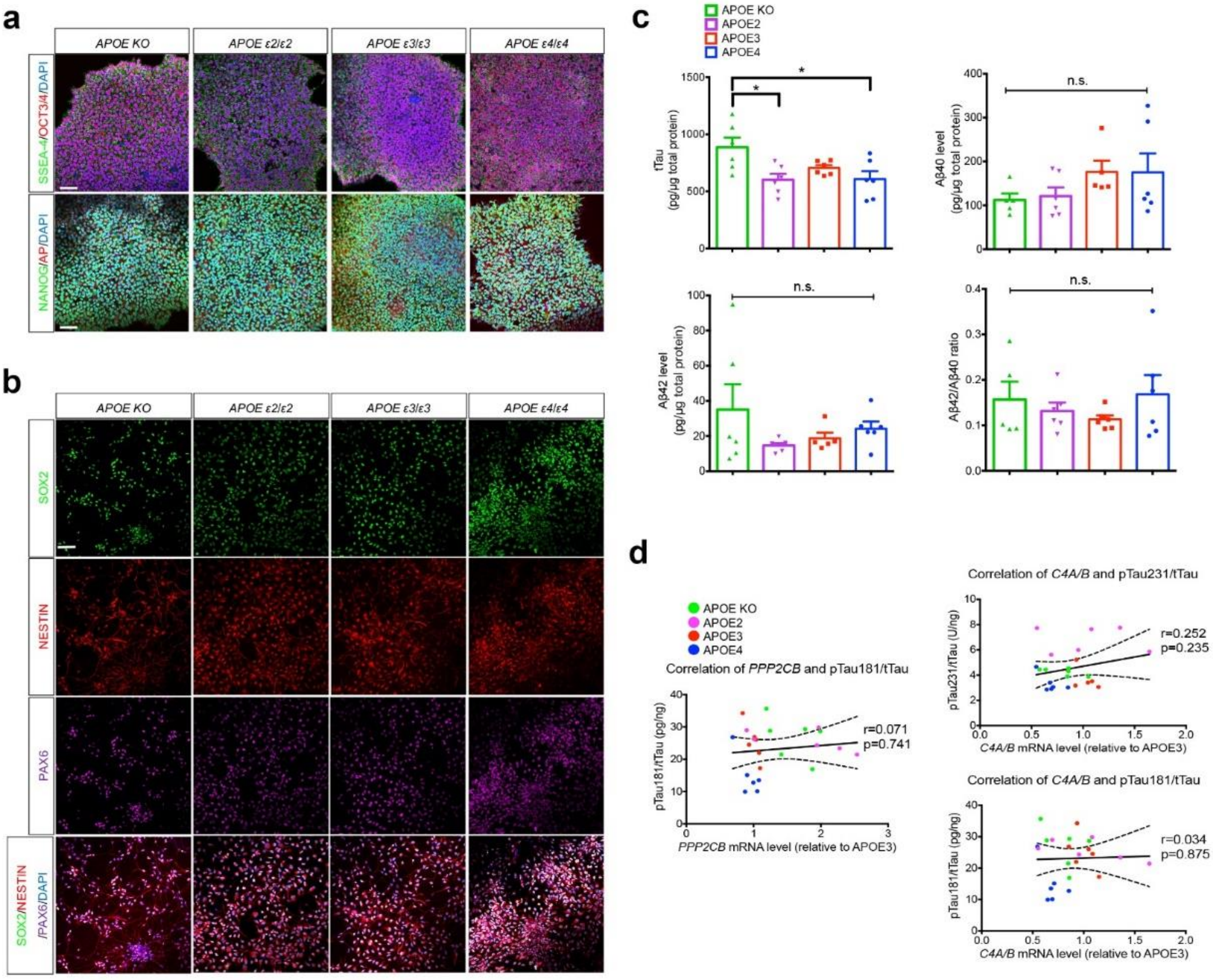

**Table S1.** Sample size and genotyping platform of the ADGC cohorts in *APOE* genotypes

| Cohort | Platform | ε2/3+ε2/3 | | ε3/3 | | ε2/4 | | ε3/4 | | ε4/4 | |
|---|---|---|---|---|---|---|---|---|---|---|---|
| | | Cases | Controls | Cases | Controls | Cases | Controls | Cases | Controls | Cases | Controls |
| ACT1 | Illumina 660 | 36 | 193 | 249 | 955 | 17 | 25 | 181 | 276 | 36 | 14 |
| ACT2 | Illumina OmniExpress | 2 | 1 | 9 | 3 | 2 | 0 | 7 | 3 | 1 | 0 |
| ADC1 | Ilumina 660 | 44 | 70 | 448 | 280 | 37 | 7 | 738 | 138 | 252 | 5 |
| ADC2 | Ilumina 660 | 23 | 18 | 181 | 84 | 12 | 7 | 255 | 29 | 86 | 5 |
| ADC3 | Illumina OmniExpress | 22 | 73 | 207 | 333 | 29 | 9 | 283 | 108 | 79 | 12 |
| ADC4 | Illumina OmniExpress | 11 | 50 | 118 | 205 | 9 | 10 | 119 | 78 | 32 | 12 |
| ADC5 | Illumina OmniExpress | 11 | 65 | 83 | 307 | 12 | 12 | 131 | 95 | 35 | 12 |
| ADC6 | Illumina OmniExpress | 11 | 36 | 75 | 199 | 4 | 5 | 94 | 82 | 22 | 5 |
| ADC7 | Illumina OmniExpressExome | 26 | 74 | 163 | 458 | 17 | 19 | 227 | 221 | 77 | 13 |
| ADNI | Illumina 610 | 6 | 21 | 80 | 108 | 8 | 4 | 130 | 36 | 44 | 4 |
| BIOCARD | Illumina OmniExpress | 0 | 14 | 3 | 61 | 1 | 3 | 1 | 29 | 1 | 4 |
| CHAP2 | Illumina OmniExpress | 4 | 16 | 15 | 91 | 1 | 5 | 5 | 28 | 2 | 3 |
| EAS | Illumina OmniExpress | 0 | 18 | 7 | 89 | 0 | 8 | 2 | 25 | 0 | 1 |
| GenADA | Affymetrix 500/Illumina 550/ Illumina 1M | 29 | 93 | 203 | 449 | 23 | 18 | 313 | 141 | 84 | 11 |
| RMAYO | Illumina OmniExpress | 18 | 133 | 205 | 601 | 15 | 28 | 298 | 243 | 110 | 19 |
| MIRAGE | Illumina 610/ Illumina 330 | 24 | 65 | 178 | 371 | 16 | 14 | 192 | 217 | 60 | 44 |
| NBB | Illumina1M | 4 | 5 | 17 | 20 | 0 | 0 | 25 | 5 | 5 | 0 |
| NIA-LOAD | Illumina 610 | 43 | 176 | 405 | 814 | 38 | 48 | 970 | 472 | 318 | 52 |
| OHSU | Illumina HumanCNV370v1_C | 10 | 27 | 64 | 100 | 8 | 2 | 45 | 21 | 4 | 1 |
| PFIZER | Illumina 500/610 | 31 | 105 | 190 | 497 | 17 | 15 | 361 | 131 | 97 | 14 |
| RMAYO | Illumina OmniExpress | 3 | 22 | 3 | 145 | 1 | 5 | 3 | 41 | 2 | 1 |
| ROSMAP1 | Affymetrix 6.0 | 22 | 114 | 149 | 419 | 4 | 8 | 100 | 98 | 4 | 11 |
| ROSMAP2 | Illumina OmniExpress | 5 | 13 | 26 | 45 | 2 | 0 | 15 | 14 | 1 | 1 |
| TARC1 | Affymetrix 6.0 | 8 | 23 | 106 | 110 | 10 | 3 | 140 | 39 | 41 | 5 |
| TGEN2 | Affymetrix 1M | 20 | 53 | 195 | 227 | 24 | 7 | 274 | 62 | 99 | 7 |
| UKS | Illumina 550 | 37 | 1 | 225 | 5 | 19 | 0 | 233 | 1 | 77 | 0 |
| UM/VU/MSSM1 | Illumina Human550/610-Quad | 15 | 83 | 152 | 397 | 12 | 12 | 210 | 143 | 79 | 13 |
| UM/VU/MSSM2 | Illumina 1M/1M-Duo | 22 | 43 | 173 | 229 | 7 | 5 | 131 | 52 | 31 | 4 |
| UM/VU/MSSM3 | Affymetrix 6.0 | 5 | 18 | 93 | 87 | 4 | 2 | 139 | 21 | 54 | 1 |
| UM/VU/TARC2 | Illumina OmniExpress | 6 | 9 | 84 | 88 | 10 | 1 | 103 | 20 | 28 | 1 |
| UPITT | Ilumina Omni-quad | 43 | 117 | 445 | 543 | 37 | 17 | 523 | 137 | 110 | 9 |
| WASHU1 | Illumina Human610 | 24 | 23 | 117 | 113 | 13 | 2 | 130 | 43 | 30 | 5 |
| WASHU2 | Illumina OmniExpress | 3 | 5 | 13 | 50 | 2 | 1 | 16 | 13 | 3 | 2 |
| WHICAP | Illumina OmniExpress | 6 | 70 | 51 | 371 | 1 | 7 | 14 | 102 | 1 | 7 |

ACT: Adult Changes in Thought Study; ADCs: the National Institute on Aging (NIA) Alzheimer Disease Centers; ADNI: Alzheimer Disease Neuroimaging Initiative; BIOCARD: Biomarkers of Cognitive Decline Among Normal Individuals; CHAP: Chicago Health and Aging Project; EAS: Einstein Aging Study; GenADA: Multi-Site Collaborative Study for GenotypePhenotype Associations in Alzheimers Disease; MAYO: Mayo Clinic; MIRAGE: Multi-Institutional Research on Alzheimer's Genetic Epidemiology; NIA-LOAD: NIA Late-Onset Alzheimer's Disease Family Study; NBB: Netherlands Brain Bank; OHSU: Oregon Health and Science University; RMAYO: Pfizer, Rochester Mayo Clinic; ROSMAP: Rush University Religious Orders Study/Memory and Aging Project; TARCC: Texas Alzheimer's Research and Care Consortium; TGEN2: Translational Genomics Research Institute series 2; University of Miami/Vanderbilt University University/Mt. Sinai School of Medicine; UKS: Universitatsklinikum Saarlandes; UPITT: University of Pittsburgh; WASHU: Washington University; WHICAP: Washington Heights-Inwood Community Aging Project

**Table S2.** Phenotype characteristics from the ADGC cohorts in *APOE* subgroups

| *APOE* Genotype | *APOE* Subgroup | # of Eligible Cohorts | Total N | Cases | | | Control | | |
|---|---|---|---|---|---|---|---|---|---|
| | | | | N | AAO (Mean±SD) | % Female | N | AAE (Mean±SD) | % Female |
| ε2/2+ ε2/3+ ε3/3+ ε2/4+ ε3/4+ ε4/4 | ALL | 29 | 28,161 | 13,909 | 74.2±7.4 | 59 | 14,206 | 76.5±8.1 | 60 |
| ε2/2+ ε2/3 | E2P | 20 | 2,096 | 485 | 77.5±8.2 | 55 | 1,611 | 77.6±8.3 | 60 |
| ε3/3 | E33 | 29 | 13,036 | 4,485 | 76.4±7.9 | 60 | 8,551 | 77.0±8.1 | 60 |
| ε2/4 | NA | 10 | 412 | 204 | 74.9±6.5 | 65 | 208 | 75.1±7.8 | 56 |
| ε3/4 | E34 | 27 | 9,164 | 6,132 | 73.5±6.6 | 60 | 3,032 | 74.7±7.8 | 60 |
| ε4/4 | E44 | 11 | 1,223 | 1,007 | 70.3±5.8 | 57 | 216 | 72.0±6.9 | 63 |
| ε2/2+ ε2/3+ ε3/3 | E2INT | 29 | 15,359 | 5,017 | 76.5±7.9 | 60 | 10,342 | 77.1±8.2 | 60 |
| ε3/3+ ε3/4+ ε4/4 | E4INT | 29 | 24,379 | 12,471 | 74.0±7.3 | 59 | 11,908 | 76.3±8.1 | 60 |

### of Eligible Studies: The number of studies in ADGC used for a genome-wide association (GWA) analysis in an APOE subgroup. We conducted a GWA analysis when a study contained at least 10 subjects in both cases and controls and then meta-analyzed.
NA: not applicable for genetic analysis since we excluded ε2/4 carriers in APOE stratified GWA analysis but the entire sample (ALL) included ε2/4 carriers
AAO: Estimated age at dementia onset (when available) in the Alzheimer's dementia cases in years.
AAE: Age at last clinical evaluation (when available) in the combined cases and controls in years.
Mean (Mean) and standard deviation (SD) for AAO, AAD, and AAE ranged from Mean-SD and Mean+SD, Mean±SD.

**Table S3.** Significant SNP associations with AD ($P<10^{-6}$) among persons lacking *APOE* ε4

| Subgroup | SNP | Near Gene | CH | A1 | A2 | Freq1 | E2P | | | E33 | | | E2INT | | |
|---|---|---|---|---|---|---|---|---|---|---|---|---|---|---|---|
| | | | | | | | OR | 95% CI | P | OR | 95% CI | P | OR | 95% CI | P |
| E2P | rs77907104 | *LOC105374313* | 3 | C | T | 0.02 | 8.67 | 4.29 - 17.53 | **1.7E-09** | 1.10 | 0.77 - 1.55 | 6.1E-01 | NA | NA | NA |
| E2P | rs117296832 | *PPP2CB* | 8 | A | G | 0.03 | 3.94 | 2.37 - 6.54 | 1.1E-07 | 1.12 | 0.95 - 1.32 | 1.6E-01 | 2.46 | 1.56 - 3.86 | 9.8E-05 |
| E2P | rs78802006 | *KDM4C* | 9 | C | G | 0.04 | 3.79 | 2.25 - 6.40 | 6.0E-07 | 1.14 | 0.94 - 1.39 | 1.8E-01 | 2.66 | 1.53 - 4.63 | 5.6E-04 |
| E2P | rs17038845 | *RIC8B* | 12 | A | G | 0.03 | 3.55 | 2.16 - 5.86 | 6.7E-07 | 1.20 | 1.00 - 1.45 | 5.1E-02 | 2.55 | 1.58 - 4.13 | 1.3E-04 |
| E2P | rs57056064 | *RBFOX1* | 16 | C | T | 0.06 | 2.34 | 1.67 - 3.28 | 7.7E-07 | 1.07 | 0.94 - 1.20 | 3.1E-01 | 1.88 | 1.37 - 2.57 | 7.8E-05 |
| E2P | rs76084405 | *STAT5B* | 17 | A | G | 0.03 | 4.84 | 2.68 - 8.72 | 1.6E-07 | 1.05 | 0.84 - 1.31 | 6.6E-01 | 4.27 | 2.33 - 7.80 | 2.5E-06 |
| E33 | rs17447007 | *PTK2B* | 8 | G | A | 0.34 | 1.08 | 0.9 - 1.29 | 4.0E-01 | 1.18 | 1.11 - 1.25 | **3.8E-08** | 0.91 | 0.78 - 1.07 | 2.7E-01 |
| E33 | rs34173062 | *SHARPIN* | 8 | A | G | 0.08 | 1.25 | 0.82 - 1.91 | 3.0E-01 | 1.45 | 1.25 - 1.67 | 6.3E-07 | 1.00 | 0.68 - 1.46 | 9.8E-01 |
| E33 | rs7920721 | *USP6NL* | 10 | G | A | 0.39 | 1.07 | 0.90 - 1.28 | 4.4E-01 | 1.18 | 1.11 - 1.25 | **3.9E-08** | 0.93 | 0.79 - 1.10 | 3.9E-01 |
| E33 | rs1960350 | *LINC01499* | 11 | T | C | 0.22 | 1.09 | 0.87 - 1.36 | 4.6E-01 | 1.19 | 1.12 - 1.28 | 3.4E-07 | 0.89 | 0.73 - 1.08 | 2.4E-01 |
| E33 | rs79279100 | *TSPOAP1-RNF43* | 17 | G | T | 0.04 | 1.64 | 1.02 - 2.62 | 3.9E-02 | 0.59 | 0.49 - 0.71 | **2.6E-08** | 2.31 | 1.47 - 3.63 | 3.0E-04 |
| E33 | rs62084370 | *TRIM37* | 17 | A | G | 0.04 | 1.69 | 1.03 - 2.75 | 3.6E-02 | 0.58 | 0.48 - 0.70 | **1.5E-08** | 2.40 | 1.50 - 3.85 | 2.6E-04 |
| E33 | rs3865444 | *CD33* | 19 | A | C | 0.30 | 0.98 | 0.81 - 1.17 | 7.9E-01 | 0.85 | 0.80 - 0.91 | 6.9E-07 | 1.23 | 1.04 - 1.45 | 1.7E-02 |
| E2INT,E2P | rs77786537 | *RIPOR2* | 6 | G | A | 0.11 | 1.97 | 1.51 - 2.56 | 5.8E-07 | 0.94 | 0.86 - 1.03 | 1.9E-01 | 1.94 | 1.52 - 2.49 | 1.1E-07 |
| E2INT | rs62239974 | *CTDSPL* | 3 | A | C | 0.05 | 2.64 | 1.70 - 4.09 | 1.4E-05 | 0.80 | 0.69 - 0.93 | 4.3E-03 | 2.70 | 1.82 – 4.00 | 7.0E-07 |
| E2INT | rs75737704 | *FBXW7* | 4 | A | G | 0.02 | 2.05 | 0.94 - 4.49 | 7.2E-02 | 1.04 | 0.82 - 1.33 | 7.5E-01 | 5.85 | 3.06 - 11.2 | 9.8E-08 |
| E2INT | rs77005801 | *HTR1A* | 5 | G | A | 0.04 | 2.86 | 1.79 - 4.56 | 1.1E-05 | 0.96 | 0.81 - 1.12 | 5.9E-01 | 2.89 | 1.89 - 4.40 | 8.5E-07 |
| E2INT | rs9478555 | *CNKSR3* | 6 | A | G | 0.17 | 1.53 | 1.22 - 1.92 | 2.4E-04 | 0.89 | 0.83 - 0.97 | 6.0E-03 | 1.70 | 1.38 - 2.08 | 4.6E-07 |
| E2INT | rs77454027 | *ETV1* | 7 | C | T | 0.02 | 3.13 | 1.58 - 6.19 | 1.0E-03 | 0.92 | 0.73 - 1.16 | 4.9E-01 | 5.38 | 2.98 - 9.73 | **2.5E-08** |
| E2INT | rs6485360 | *HNRNPKP3* | 11 | A | G | 0.03 | 2.70 | 1.72 - 4.24 | 1.6E-05 | 0.94 | 0.78 - 1.12 | 4.6E-01 | 2.93 | 1.91 - 4.51 | 9.6E-07 |
| E2INT | rs17239735 | *CGNL1* | 15 | C | T | 0.19 | 1.72 | 1.37 - 2.16 | 3.8E-06 | 0.94 | 0.87 - 1.01 | 8.0E-02 | 1.83 | 1.49 - 2.24 | **4.8E-09** |
| E2INT | rs118165364 | *GALR1-LINC01029* | 18 | G | A | 0.02 | 2.15 | 1.02 - 4.55 | 4.5E-02 | 1.10 | 0.88 - 1.37 | 4.2E-01 | 4.52 | 2.49 - 8.22 | 7.4E-07 |

SNPs were included if the heterogeneity degree of freedom (HetDf) in the meta-analysis containing at least half of eligible datasets for the given subgroup.
Genome-wide significant results ($P<5x10^{-8}$) are highlighted in bold.
Odds ratio (OR), 95% confidence interval (95% CI), and P-value (P) were calculated using a logistic regression model including covariates for age, sex, and PCs.

**Table S4.** Significant associations with AD ($P<10^{-6}$) among persons lacking *APOE* ε2

| Subgroup | SNP | Near Gene | CH | A1 | A2 | Freq1 | E34 | | | E44 | | | E4INT | | |
|---|---|---|---|---|---|---|---|---|---|---|---|---|---|---|---|
| | | | | | | | **OR** | **95% CI** | **P** | **OR** | **95% CI** | **P** | **OR** | **95% CI** | **P** |
| E34 | rs4663105 | *BIN1* | 2 | C | A | 0.44 | 1.28 | 1.19 - 1.38 | **4.9E-11** | 0.93 | 0.73 - 1.2 | 6.0E-01 | 1.05 | 0.97 - 1.13 | 2.6E-01 |
| E34 | rs1603122 | *NPTX2* | 7 | C | G | 0.07 | 0.70 | 0.61 - 0.8 | 2.3E-07 | 1.16 | 0.71 - 1.91 | 5.5E-01 | 0.85 | 0.74 - 0.98 | 2.8E-02 |
| E34 | rs4766794 | *MAP1LC3B2* | 12 | C | T | 0.02 | 0.49 | 0.38 - 0.64 | 6.7E-08 | 0.62 | 0.24 - 1.58 | 3.2E-01 | 0.64 | 0.5 - 0.83 | 6.3E-04 |
| E44 | rs75983775 | *MIR3681HG-TRIB2* | 2 | T | G | 0.02 | 1.16 | 0.84 - 1.61 | 3.6E-01 | 0.12 | 0.07 - 0.23 | **9.4E-11** | 0.69 | 0.49 - 0.97 | 3.0E-02 |
| E44 | rs115637942 | *KLHL29* | 2 | A | G | 0.02 | 1.14 | 0.82 - 1.59 | 4.3E-01 | 0.13 | 0.06 - 0.27 | **1.8E-08** | 0.65 | 0.47 - 0.9 | 8.8E-03 |
| E44 | rs79385707 | *ALK* | 2 | T | G | 0.04 | 0.98 | 0.83 - 1.16 | 8.0E-01 | 0.23 | 0.14 - 0.4 | 1.3E-07 | 1.12 | 0.93 - 1.35 | 2.2E-01 |
| E44 | rs180795945 | *CTNNA2* | 2 | T | C | 0.01 | NA | NA | NA | 0.06 | 0.02 - 0.15 | **6.1E-09** | NA | NA | NA |
| E44 | rs139471546 | *RPRM-GALNT13* | 2 | G | A | 0.03 | 0.99 | 0.77 - 1.27 | 9.2E-01 | 0.19 | 0.1 - 0.36 | 1.9E-07 | 0.69 | 0.53 - 0.89 | 4.9E-03 |
| E44 | rs56112638 | *PLEKHM3* | 2 | A | C | 0.35 | 0.99 | 0.92 - 1.07 | 8.0E-01 | 0.53 | 0.41 - 0.68 | 9.2E-07 | 0.93 | 0.86 - 1.01 | 8.0E-02 |
| E44 | rs113369806 | *TENM3* | 4 | T | G | 0.06 | 1.15 | 0.98 - 1.35 | 9.6E-02 | 0.30 | 0.19 - 0.48 | 4.3E-07 | 0.90 | 0.76 - 1.07 | 2.4E-01 |
| E44 | rs117768391 | *HECW1* | 7 | A | G | 0.04 | 0.91 | 0.72 - 1.15 | 4.3E-01 | 0.23 | 0.13 - 0.4 | 3.8E-07 | 0.73 | 0.59 - 0.91 | 4.8E-03 |
| E44 | rs143843879 | *DNAJB5* | 9 | A | G | 0.02 | 1.03 | 0.71 - 1.51 | 8.6E-01 | 0.07 | 0.03 - 0.19 | 1.8E-07 | 0.57 | 0.39 - 0.83 | 3.1E-03 |
| E44 | rs78277278 | *FAM92B* | 16 | T | C | 0.01 | 0.94 | 0.65 - 1.36 | 7.4E-01 | 0.09 | 0.04 - 0.22 | 1.0E-07 | 0.96 | 0.63 - 1.46 | 8.3E-01 |
| E44 | rs139527441 | *ZNF443* | 19 | A | T | 0.02 | 0.90 | 0.64 - 1.26 | 5.3E-01 | 0.10 | 0.04 - 0.21 | **9.2E-09** | 0.70 | 0.48 - 1.03 | 7.2E-02 |
| E4INT | rs76765792 | *NKX2-3* | 10 | T | C | 0.01 | 0.44 | 0.32 - 0.62 | 1.9E-06 | NA | NA | NA | 0.37 | 0.26 - 0.52 | **4.0E-08** |
| E4INT | rs139641486 | *ARNT2* | 15 | A | G | 0.02 | 0.50 | 0.36 - 0.7 | 3.9E-05 | NA | NA | NA | 0.45 | 0.33 - 0.62 | 7.2E-07 |

SNPs were included if the heterogeneity degree of freedom (HetDf) in the meta-analysis containing at least half of eligible datasets for the given subgroup.
Genome-wide significant results ($P<5x10^{-8}$) are highlighted in bold.
Odds ratio (OR), 95% confidence interval (95% CI), and P-value (P) were calculated using a logistic regression model including covariates for age, sex, and PCs.

**Table S5.** Biologically connected networks including genes significantly associated with AD in *APOE* genotype stratified GWAS

| Network.ID | Molecules in Network | Score | # of GWA genes |
|---|---|---|---|
| E2INT.1 | **ADGRV1**,AKT1,APC,**ASAP1**,BCL6,CALCA,CDH1,**CGNL1**,**DAAM1**,**DNAH9**,**ECE1**,EED,EEF1G,**EMSY**,HMMR,HSPA5,IFNAR2,IL5,MYO, NFkB complex,PLTP,PTH,**RIPOR2**,**SPATA17**,**ST3GAL1**,**STAT5B**,TGFB1,TJP1,TRAF7,VCL, WNK1,YWHA, YWHAG,YWHAZ | 30 | 11 |
| E2INT.2 | **ADAMTS14**,AKT1,APC,**AVPR1A**,CALCA,CDK2,**CNKSR3**,**CTDSPL**,CTNNB1,**DNAJC1**,EED,EEF1G,EGFR,**EIPR1**,**GRID1**,HMMR,**HOMER2**,HSPA5,MAP2K2,MYOF,**PCDH7**,PDIA5,PTH,RAC1,RCN2,**SRGAP3**,TJP1,TNF,TP53,USP31,VCL,WNT3A,YWHAE,YWHAG,YWHAZ | 24 | 10 |
| E2P.1 | **ARHGAP32**,ATR,AXIN1,CAV1,CCNA2,CCT5,CDK1,**DNTTIP1**,ETS2,histone h3,INTS1,INTS3,INTS5,INTS6,**ITGA1**,KCNG1,**KDM4C**, KLK3,MIR205HG,PCNX2,**PMEPA1**,POLR2A,**PPP2CB**,PP2R1A,PPP2R1B,PRKCD,**PTPRT**,SSPN,**STAT3**,STRN3,TCP1,TP53,**TP63**,TRMT11, **UBE2C** | 21 | 10 |
| E2P.2 | **CCL16**,**CSGALNACT1**,CXCL6,EGF,EGFR,EPO,EZH2,FER,FOXP3,GNA15,**GPR63**,GH1,HAMP,IL10,IL15,IL2,IL21,IL22RA2,IL31,IL7,IL9, JAK2,**KCNK1**,LILRB2,MAF,MHC Class II complex,NFATC2,PDCD1LG2,PGR,PPARG,PPIF,PRL,SRC,**STAT3**,**STAT5B** | 11 | 6 |
| E33.1 | **ABCA7**,APP,**ARRDC4**,**ATE1**,**CD33**,CD63,CDH1,**CYCS**,DDR1,**ECM1**,EZH2,**FALEC**,GSK3B,IL13,IL24,IL36A,JPH3,**KIAA0513**,**MCL1**, **MS4A4A**,NFkB complex,PARP9,PAWR,PI3K complex,**PRUNE**,**PTK2B**,**RAD51C**,RASSF5,RASSF9,SELENON, **SHARPIN**,SMARCA4,TGM2,**TRIM37**,WNT10B | 37 | 15 |
| E34.1 | ALB,AMER1,APP,**AXIN2**,**BIN1**,C1q,C5,**CCR6**,**CDH13**,COL18A1,**CR1**, CXCL8,**ELL**,Estrogen receptor,GSK3A,**MAP1LC3B2**,mir-22-3p, MYC,**NEFL**,**NEFM**,NOTCH1,NOTCH2,NR3C1,NUPR1,**PDGFD**,RB1,TCP10,**TCP10L2**,TGFB1,TGM2,TNF, **TNS3**,TP53,TWIST2,WNT5A | 25 | 12 |
| E34.2 | AGTR1,**AP3B2**,BCL6,CD101,CD28,CD3,CD40,CTNNB1,EGFR,ELAVL1,GSK3B,**HOMER2**,IFNG,IGM,IL10,IL13,IL1B,IL2,IL27,IL3,Interferon alpha,KITLG,**MS4A2**,MS4A3,**MS4A4A**,MYC,PRMT1,**RNF213**,**SAMSN1**,STAT1,TGFB1,TGM2,TNF,TP53,**TRPV4** | 13 | 7 |
| E44.1 | **ADGRL2**,Akt,**ALK**,Ap1,**BIN1**,**CACNA1A**,**CNBP**,**CTNNA2**,**CYTH1**,ERK1/2,G protein beta gamma complex,**HECW1**,Jnk,**KLHL29**,**LHPP**, **LINC00467**,MAP2K1,MAP2K2,**MCC**,NCOR1,NFkB complex,**NR3C1**,P38 MAPK,p70 S6k,PI3K complex,Pka,**PRKAA**,PRKAA1,**RAB43**, **SCD5**,**SMC2**,**ST6GAL1**,TACR1,Vegf,**YAP1** | 51 | 19 |
| E44.2 | **AGAP1**,ATP5F1B,BCL6,CAV1,CDCA3,EPHA2,G3BP1,GAN,GLUD1,HOOK1,HSPA9,HSPD1,IL5,IRF5,ITGA1,JUP,LRP1,MCM2,**PABPC4L**, **PLEKHM3**,PPIA,PPP2R1A,PSEN1,RANGRF,SDHA,**SGCZ**,SLC1A5,**SLF2**,**SPAST**,**TASP1**,**TENM3**,**THSD7B**,VIRMA,ZBTB21,**ZNF443** | 21 | 10 |
| E44.3 | ABCA2,AGO2,ANGPTL2,APC,CDH3,CDH5,CSNK1D,**DLG2**,DLGAP1,DNAJB12,**DNAJB5**,EPHA2,ESR1,F Actin,FBXW11,G3BP1,HNRNPL, HSPA5,HSPA9,IL5,IRF5,JUP,**KCNH5**,**KCNN2**,KLHL9,**LIN9**,LRP1,**LRP1B**,NUDCD3,OBSCN,PAN2,PSEN1,SSX2IP,TECPR2,TP53 | 11 | 6 |
| E4INT.1 | AQP4,Ca2,CALB2,**CCDC18**,CD59,**CERS2**,CMKLR1,CREB1,CXCR5,**DYRK1B**,**FAM120B**,FHIT,**GPC6**,IL6R,**KCNQ3**,LAMB3,LCAT,LRRFIP1,MAPK1,MYC,PGK1,PLA2G5,**PYY**,**RALYL**,RELA,SETDB1,SLC12A3,SLC2A3,SP1,SRC,SYT9,TNFRSF9,TRB,ZFP36L1 | 25 | 8 |

NFkB complex genes: NEK4, NFKB1, NFKB2, RELA, RELB, and REL; Histone h3: HIST3H3; Growth hormone: GH1; MHC Class II complex: HLA-A, HLA-B, HLA-C, HLA-DRB1, HLA-DQA1, and HLA-DQB1; PI3K complex: PIK3CD, PIK3CA, PIK3CB, PIK3CG, and PIK3R1; C1q: C1QA, C1QB, C1QC, C1R, and C1S; Estrogen receptor: EGFR, ESR1, and ESR2; mir-22-3p: MIR22; Interferon alpha: IFNA1; ERK1/2 genes: MAPK1 and MAPK3; G protein beta gamma complex: GNB1, and GNGT1; Jnk gene: MAPK8, MAPK9, and MAPK10; P38 MAPK gene: MAPK11, MAPK12, MAPK13, and MAPK14; p70 S6k gene: RPS6KB1; Pka: PRKACA; Vegf: VEGFA, VEGFB, VEGFC, and VEGFD; F Actin: ACTB

A total of 111 unique genes (highlighted in bold) genes containing a SNP significantly associated with AD ($P<10^{-5}$) in the corresponding *APOE* subgroup were included in the network analyses.

**Table S7.** Pathway and enrichment analysis of *APOE* related networks

| Network.ID | Neuropath Associated Gene Enrichment P-value | | Top Ranked Pathways with FDR P<0.05 from the Reactome Database |
|---|---|---|---|
| | Plaques | Tangles | |
| E2INT.1 | 7.8E-02 | 8.4E-02 | Cytokine signaling in immune system; Signaling by interleukins; Diseases of signal transduction by growth factor receptors; Interleukins 2, 3, and 5 and GM-CSF signaling |
| E2INT.2 | 1.0E-01 | 1.1E-01 | Inactivation of CDC42 and RAC1 |
| E2P.1 | 2.8E-03 | 7.7E-05 | RNA polymerase II transcription; Cellular responses to stress; Regulation of TP53 expression and degradation; Signaling by Rho GTPases; Signaling by WNT and receptor tyrosine kinases; Senescence-associated secretory phenotype (ASAP); Chaperonin-mediated protein folding; Apoptosis; Mitotic spindle checkpoint |
| E2P.2 | 1.0E+00 | 1.0E+00 | Cytokine signaling in immune system; Growth hormone receptor signaling; Interleukin signaling |
| E33.1 | 1.0E+00 | 1.2E-01 | Cytokine signaling in immune system; Interleukin signaling; Adaptive immune system; Innate immune system |
| E34.1 | 4.1E-03 | 4.8E-03 | TCF dependent signaling in response to WNT; Beta-catenin independent WNT signaling; Deubiquitination; Regulation of complement cascade |
| E34.2 | 6.3E-02 | 6.8E-02 | NA |
| E44.1 | 1.1E-02 | 1.3E-02 | Signaling by receptor tyrosine kinases; Nuclear receptor transcription pathway; Nervous system development; RNA polymerase II transcription; RUNX2 regulates osteoblast differentiation; Signaling by ERBB4; Infectious disease; EGR2 and SOX10-mediated initiation of Schwann cell myelination |
| E44.2 | 1.0E+00 | 1.0E+00 | NA |
| E44.3 | 9.7E-04 | 5.2E-02 | NA |
| E4INT.1 | 1.0E+00 | 5.2E-02 | NA |

Neuropath Associated Gene Enrichment P-value: P value from enrichment analysis in Fisher's exact tests using significant genes with $P<10^{-3}$ from Beecham et al. 2014.[53] NA: Significant pathways were not available at FDR P<0.05.

**Table S8.** Characteristics of neuropathologically examined brain samples by cohort and *APOE* genotype

| APOE Subgroup | ROSMAP | | | | | | | FHS/BUADC | | | | |
|---|---|---|---|---|---|---|---|---|---|---|---|---|
| | AD | Control | RNAseq | snRNAseq | mQT | Plaques* | Tangles* | AD | Control | RNAseq | Plaques* | Tangles* |
| E23 | 32 | 39 | 71 | 11 | 92 | 0.55 ± 0.9 | -0.43 ± 0.8 | 6 | 31 | 37 | 0.52 ± 0.8 | -0.55 ± 0.7 |
| E33 | 193 | 156 | 349 | 23 | 413 | 0.18 ± 1.0 | -0.20 ± 1.0 | 33 | 76 | 109 | 0.13 ± 1.0 | -0.09 ± 1.0 |
| E34 | 101 | 24 | 125 | 9 | 162 | -0.45 ± 0.9 | 0.36 ± 0.9 | 19 | 12 | 31 | -0.45 ± 0.9 | 0.47 ± 1.0 |
| ALL | 339 | 229 | 568 | 48 | 697 | 0.08 ± 1.0 | -0.10 ± 1.0 | 64 | 129 | 193 | 0.06 ± 1.0 | -0.03 ± 1.0 |

RNAseq: bulk RNA sequencing data; snRNAseq: single nuclei RNA sequencing data; mQT: array data of methylation levels of CpG sites. Numbers of AD cases and controls, and subjects with RNA sequence (RNAseq), single nuclei RNA sequence (snRNAseq, and quantified methylation data are shown.

***** Mean ± standard deviation

**Table S9.** Association of expression levels of top ranked genes in *APOE* ε2 group-related networks with plaque and tangle density

| Network | Gene | AD vs. Control | | | Plaque density | | | | Tangle density | | | |
|---|---|---|---|---|---|---|---|---|---|---|---|---|
| | | **Z** | **P-value** | **DIR** | **β** | **SE** | **P-value** | **DIR** | **β** | **SE** | **P-value** | **DIR** |
| E2INT.1 | *ASAP1* | -1.41 | 1.6E-01 | -- | 0.03 | 0.03 | 3.7E-01 | ++ | -0.07 | 0.03 | 2.1E-02 | -- |
| E2INT.1 | *CGNL1* | 5.06 | **4.2E-07** | ++ | -0.06 | 0.04 | 1.7E-01 | -- | 0.10 | 0.04 | 2.1E-02 | ++ |
| E2INT.1 | *DNAH9* | -2.13 | 3.3E-02 | -- | 0.13 | 0.06 | 2.8E-02 | ++ | -0.15 | 0.05 | 7.2E-03 | -- |
| E2INT.1 | *ECE1* | 6.84 | **8.1E-12** | ++ | -0.09 | 0.04 | 4.7E-02 | -- | 0.08 | 0.04 | 4.6E-02 | ++ |
| E2INT.1 | *RIPOR2* | -2.18 | 2.9E-02 | -- | 0.04 | 0.04 | 3.3E-01 | -+ | -0.07 | 0.04 | 5.7E-02 | -- |
| E2INT.1 | *ST3GAL1* | 2.1 | 3.6E-02 | ++ | 0.07 | 0.04 | 6.5E-02 | ++ | -0.06 | 0.03 | 8.8E-02 | -- |
| E2INT.1; E2P.2 | *STAT5B* | 3.46 | **5.4E-04** | ++ | -0.01 | 0.03 | 7.4E-01 | -- | 0.00 | 0.03 | 9.8E-01 | +- |
| E2INT.2 | *AVPR1A* | 2.19 | 2.9E-02 | ++ | -0.03 | 0.04 | 4.4E-01 | -+ | -0.02 | 0.03 | 5.4E-01 | +- |
| E2INT.2 | *CNKSR3* | 3.97 | **7.2E-05** | ++ | -0.12 | 0.05 | 1.1E-02 | -- | 0.15 | 0.04 | **1.1E-03** | ++ |
| E2INT.2 | *CTDSPL* | 4.62 | **3.9E-06** | ++ | -0.03 | 0.03 | 3.7E-01 | -- | 0.02 | 0.03 | 5.3E-01 | ++ |
| E2INT.2 | *DNAJC1* | 5.05 | **4.4E-07** | ++ | -0.05 | 0.04 | 2.2E-01 | -- | 0.02 | 0.04 | 5.7E-01 | -+ |
| E2INT.2 | *EIPR1* | -3.23 | **1.3E-03** | -- | 0.08 | 0.05 | 1.0E-01 | ++ | -0.13 | 0.04 | **1.4E-03** | -- |
| E2INT.2 | *GRID1* | 2.06 | 4.0E-02 | -+ | -0.05 | 0.05 | 3.0E-01 | -- | -0.01 | 0.04 | 8.3E-01 | +- |
| E2P.1 | *ARHGAP32* | -2.6 | 9.4E-03 | -- | 0.05 | 0.05 | 2.9E-01 | -+ | -0.13 | 0.04 | 4.1E-03 | +- |
| E2P.1 | *DNTTIP1* | -2.18 | 2.9E-02 | -- | 0.05 | 0.05 | 3.2E-01 | ++ | -0.08 | 0.04 | 4.5E-02 | -- |
| E2P.1 | *ITGA1* | 2.64 | 8.4E-03 | +- | 0.00 | 0.04 | 9.4E-01 | -+ | 0.03 | 0.04 | 4.7E-01 | ++ |
| E2P.1 | *KDM4C* | 0.55 | 5.8E-01 | +- | 0.05 | 0.04 | 1.6E-01 | ++ | -0.09 | 0.03 | 1.1E-02 | -- |
| E2P.1 | *PMEPA1* | 3.29 | **9.9E-04** | ++ | -0.05 | 0.04 | 2.4E-01 | -- | 0.03 | 0.03 | 3.3E-01 | ++ |
| E2P.1 | *TP63* | 2.11 | 3.5E-02 | ++ | 0.00 | 0.03 | 9.9E-01 | +- | 0.04 | 0.03 | 1.3E-01 | -+ |
| E2P.1; E2P.2 | *STAT3* | 3.74 | **1.8E-04** | ++ | -0.02 | 0.04 | 6.1E-01 | +- | 0.02 | 0.04 | 6.0E-01 | ++ |
| E2P.2 | *CSGALNACT1* | -1.36 | 1.7E-01 | +- | 0.02 | 0.04 | 5.9E-01 | ++ | -0.08 | 0.04 | 5.0E-02 | -- |
| E2P.2 | *KCNK1* | -3.46 | **5.4E-04** | -- | 0.06 | 0.05 | 2.8E-01 | ++ | -0.05 | 0.05 | 3.6E-01 | +- |

Eexpression measured in bulk RNA sequencing (Bulk RNAseq) data; results shown derived from meta-analysis of results in the ROSMAP and FHS/BUADC datasets; Top ranked *APOE* ε2 related genes include those that were at least nominally significant (P<0.05) in tests of differential expression between AD cases and controls, plaque density (CERADscore), or tangle density (Braak stage).
Significant P-values corrected for multiple testing (threshold P=0.00135) are highlighted in bold.

**Table S10.** Differential expression in brain cell types from the top ranked *APOE* ε2 related genes

| Network.ID | Gene | Ast | | ExN | | InN | | Mic | | Oli | | OPC | |
|---|---|---|---|---|---|---|---|---|---|---|---|---|---|
| | | **LogFC** | **P** | **LogFC** | **P** | **LogFC** | **P** | **LogFC** | **P** | **LogFC** | **P** | **LogFC** | **P** |
| E2INT.1 | *ADGRV1* | -0.06 | 1.10E-02 | -0.03 | **1.2E-22** | 0.02 | 2.6E-01 | -0.15 | 3.3E-01 | 0.00 | 3.3E-01 | -0.10 | 9.4E-01 |
| E2INT.1 | *ASAP1* | 0.05 | 7.70E-01 | -0.11 | **4.5E-69** | -0.15 | **2.1E-10** | -0.08 | 9.0E-01 | -0.08 | 3.4E-01 | -0.11 | 4.3E-02 |
| E2INT.1 | *DAAM1* | 0.08 | 8.70E-01 | -0.09 | **3.4E-84** | -0.20 | **1.3E-21** | -0.04 | 8.0E-01 | -0.09 | 7.2E-01 | -0.11 | 2.4E-01 |
| E2INT.1 | *DNAH9* | -0.01 | 5.70E-01 | -0.04 | **8.2E-13** | -0.03 | 6.5E-02 | NA | NA | -0.01 | 6.4E-01 | 0.10 | 2.9E-02 |
| E2INT.1 | *ECE1* | 0.02 | 7.10E-01 | 0.02 | **4.2E-09** | 0.00 | 2.7E-03 | -0.02 | 3.0E-01 | 0.08 | 1.5E-02 | -0.08 | 9.0E-01 |
| E2INT.1 | *EMSY* | 0.01 | 5.90E-01 | -0.05 | **1.4E-31** | -0.16 | **4.6E-06** | 0.27 | 2.5E-01 | 0.05 | 5.8E-02 | 0.11 | 6.3E-01 |
| E2INT.1 | *RIPOR2* | 0.03 | 2.40E-01 | 0.03 | **7.3E-13** | -0.07 | **7.8E-08** | -0.17 | 9.3E-01 | -0.04 | 1.4E-01 | -0.12 | 2.0E-01 |
| E2INT.1 | *ST3GAL1* | -0.08 | 2.20E-01 | -0.06 | **5.9E-24** | -0.04 | 2.1E-03 | -0.37 | 1.3E-01 | 0.01 | 5.6E-01 | -0.18 | 3.0E-02 |
| E2INT.1; E2P.2 | *STAT5B* | 0.07 | 7.20E-01 | -0.04 | **1.4E-17** | 0.11 | 1.3E-01 | 0.37 | 4.9E-02 | 0.10 | 3.0E-02 | -0.06 | 6.0E-01 |
| E2INT.2 | *CNKSR3* | 0.09 | 6.00E-01 | -0.03 | **1.1E-11** | 0.01 | 3.8E-01 | -0.02 | 9.8E-01 | -0.04 | 5.5E-01 | -0.22 | 2.0E-01 |
| E2INT.2 | *CTDSPL* | 0.15 | 5.90E-01 | 0.03 | **4.2E-23** | 0.04 | 6.5E-02 | -0.31 | 4.4E-01 | 0.01 | 3.9E-01 | -0.05 | 1.4E-01 |
| E2INT.2 | *DNAJC1* | 0.10 | 7.30E-01 | -0.12 | **2.2E-62** | -0.15 | 7.8E-05 | -0.24 | 7.8E-01 | 0.05 | 3.0E-02 | -0.06 | 5.2E-01 |
| E2INT.2 | *EIPR1* | 0.16 | 3.40E-01 | -0.13 | **1.1E-64** | -0.15 | **1.7E-11** | 0.28 | 1.8E-01 | -0.10 | 1.3E-01 | -0.08 | 1.2E-01 |
| E2INT.2 | *GRID1* | -0.03 | 8.90E-01 | -0.03 | **4.1E-13** | -0.02 | 2.0E-03 | 0.25 | 1.5E-01 | 0.12 | 8.6E-03 | 0.01 | 7.8E-01 |
| E2INT.2 | *PCDH7* | 0.06 | 5.40E-01 | -0.10 | **1.0E-68** | -0.08 | **2.7E-06** | -0.64 | 1.5E-02 | -0.17 | 1.6E-01 | 0.06 | 4.6E-01 |
| E2INT.2 | *SRGAP3* | -0.28 | **7.60E-08** | -0.19 | **2.3E-139** | -0.26 | **6.9E-22** | -0.16 | 2.6E-01 | -0.12 | 2.7E-02 | -0.29 | 6.6E-04 |
| E2INT.2; E34.2 | *HOMER2* | 0.12 | 7.90E-01 | 0.04 | **5.4E-07** | 0.04 | 1.4E-04 | -0.14 | 6.9E-01 | -0.02 | 3.7E-01 | -0.01 | 9.0E-01 |
| E2P.1 | *ARHGAP32* | -0.17 | 1.50E-04 | -0.16 | **6.0E-104** | -0.13 | **4.9E-11** | 0.07 | 4.7E-01 | -0.11 | 2.2E-01 | -0.13 | 9.5E-02 |
| E2P.1 | *DNTTIP1* | 0.01 | 8.50E-01 | -0.05 | **3.3E-35** | -0.10 | 5.1E-05 | 0.60 | 1.7E-02 | 0.03 | 3.4E-01 | 0.02 | 6.1E-01 |
| E2P.1 | *KDM4C* | -0.01 | 7.80E-01 | -0.07 | **2.1E-35** | -0.01 | 7.2E-04 | 0.07 | 5.8E-01 | 0.01 | 7.4E-01 | -0.06 | 7.4E-01 |
| E2P.1 | *PMEPA1* | 0.03 | 4.70E-01 | -0.07 | **1.9E-37** | -0.12 | **8.5E-11** | -0.49 | 1.0E-01 | 0.05 | 1.7E-01 | 0.20 | 3.9E-01 |
| E2P.1 | *PPP2CB* | 0.19 | 9.30E-02 | -0.12 | **5.0E-74** | -0.12 | **6.7E-12** | 0.53 | 4.5E-02 | 0.07 | 6.9E-05 | -0.03 | 8.0E-02 |
| E2P.1 | *PTPRT* | 0.00 | 3.90E-01 | -0.11 | **3.4E-57** | -0.28 | **8.7E-20** | -0.02 | 4.5E-01 | -0.04 | 4.4E-01 | -0.21 | 3.0E-02 |
| E2P.1; E2P.2 | *STAT3* | 0.07 | 9.80E-01 | -0.10 | **3.0E-40** | -0.09 | 5.5E-05 | -0.29 | 3.2E-01 | 0.04 | 2.8E-02 | -0.22 | 5.4E-02 |
| E2P.2 | *CSGALNACT1* | -0.13 | 1.20E-03 | -0.14 | **6.9E-64** | -0.02 | 3.2E-03 | 0.20 | 3.8E-01 | -0.04 | 9.5E-01 | -0.04 | 2.7E-01 |
| E2P.2 | *GPR63* | 0.05 | 1.50E-01 | 0.02 | **1.0E-15** | -0.19 | **2.5E-10** | 0.43 | 1.5E-01 | 0.01 | 6.9E-01 | 0.16 | 2.5E-01 |
| E2P.2 | *KCNK1* | 0.06 | 7.80E-01 | -0.09 | **3.4E-62** | 0.12 | 6.9E-01 | 0.22 | 6.2E-01 | 0.05 | 8.8E-02 | 0.21 | 2.0E-01 |

Ast: astrocyte; ExN: excitatory neuron; InN: inhibitory neuron; Mic: microglia; Oli: oligodendrocytes; OPC: oligodendrocyte progenitor cells. Log2 fold change of expression levels (LogFC) and adjusted P value (Adj-P) were calculated between AD and control brains from single nuclei RNA sequencing data from ROSMAP. Significant false discovery rate (FDR) adjusted P-values are highlighted in bold.

**Table S11.** Differential expression between AD and control brain cell types from the previously known AD genes

| Gene | Ast | | ExN | | InN | | Mic | | Oli | | OPC | |
|---|---|---|---|---|---|---|---|---|---|---|---|---|
| | LogFC | P | LogFC | P | LogFC | P | LogFC | P | LogFC | P | LogFC | P |
| *APOE* | -0.315 | **7.3E-17** | 0.027 | **8.6E-06** | 0.037 | 5.4E-04 | 0.151 | 1.6E-03 | -0.137 | 3.6E-03 | -0.271 | 9.8E-03 |
| *ABCA7* | 0.039 | 9.7E-01 | 0.118 | 8.9E-01 | -0.032 | 3.1E-02 | -0.211 | 8.3E-01 | 0.093 | 5.0E-02 | -0.028 | 9.5E-02 |
| *ACE* | -0.018 | 8.2E-01 | -0.001 | **4.0E-08** | -0.012 | 1.8E-01 | -0.041 | 9.8E-01 | -0.030 | 5.4E-01 | -0.028 | 5.1E-01 |
| *ADAM10* | -0.130 | 7.5E-02 | -0.035 | **1.7E-23** | 0.044 | 3.1E-03 | 0.068 | 7.7E-01 | -0.002 | 9.7E-02 | -0.056 | 5.9E-01 |
| *ADAMTS1* | -0.013 | 7.9E-01 | -0.001 | 6.6E-01 | -0.003 | 2.9E-01 | 0.082 | 3.1E-01 | 0.093 | 1.1E-04 | -0.049 | 9.4E-01 |
| *BIN1* | -0.003 | 7.8E-02 | 0.060 | **1.6E-09** | -0.037 | **3.2E-07** | 0.009 | 5.6E-01 | 0.033 | 5.5E-03 | 0.162 | 2.5E-01 |
| *CASS4* | 0.027 | 9.1E-01 | 0.003 | 6.1E-01 | 0.005 | 1.5E-01 | -0.114 | 9.5E-01 | NA | NA | NA | NA |
| *CD2AP* | -0.022 | 5.3E-01 | -0.024 | **1.3E-15** | 0.041 | 6.3E-01 | -0.096 | 7.1E-01 | 0.002 | 7.2E-01 | 0.018 | 9.2E-01 |
| *CLU* | -0.082 | 4.4E-03 | -0.147 | **2.3E-76** | -0.192 | **8.4E-24** | -0.034 | 9.3E-01 | 0.055 | 4.4E-03 | -0.196 | 7.2E-05 |
| *CR1* | NA | NA | NA | NA | NA | NA | NA | NA | 0.105 | 2.6E-02 | -0.032 | 3.3E-01 |
| *ECHDC3* | 0.010 | 3.6E-01 | 0.004 | 1.2E-01 | 0.001 | 6.6E-01 | 0.001 | 6.9E-01 | NA | NA | 0.018 | 3.1E-01 |
| *FERMT2* | 0.026 | 4.4E-01 | -0.006 | 1.6E-01 | -0.044 | 2.5E-02 | -0.118 | 9.7E-01 | -0.033 | 7.6E-01 | -0.060 | 6.1E-01 |
| *HLA-DQB1* | 0.104 | 7.8E-01 | -0.157 | **1.3E-44** | -0.039 | 1.3E-01 | -0.434 | 2.7E-01 | -0.028 | 3.7E-01 | -0.032 | 8.9E-02 |
| *HLA-DRB1* | -0.104 | 1.8E-01 | -0.147 | **4.4E-82** | -0.056 | 3.7E-04 | -0.036 | 9.7E-01 | -0.025 | 4.9E-02 | -0.042 | 8.9E-02 |
| *INPP5D* | 0.350 | 6.7E-03 | -0.124 | **2.2E-69** | -0.009 | 2.3E-01 | NA | NA | 0.012 | 3.5E-01 | -0.012 | 6.8E-01 |
| *IQCK* | -0.021 | 2.7E-01 | -0.005 | 4.1E-02 | 0.016 | 9.7E-02 | -0.340 | 3.1E-02 | 0.077 | 8.4E-05 | -0.104 | 1.4E-01 |
| *MS4A4A* | 0.009 | 3.6E-01 | 0.000 | 7.3E-01 | NA | NA | -0.615 | 7.1E-02 | NA | NA | NA | NA |
| *NYAP1* | -0.030 | 5.3E-01 | -0.129 | **7.8E-59** | -0.133 | 8.8E-04 | 0.193 | 9.8E-01 | -0.035 | 3.1E-01 | 0.037 | 7.5E-01 |
| *OARD1* | -0.044 | 3.9E-01 | -0.112 | **7.1E-39** | -0.071 | 2.1E-03 | -0.244 | 5.8E-01 | -0.081 | 6.3E-01 | -0.198 | 8.1E-02 |
| *PICALM* | -0.164 | 1.8E-01 | -0.062 | **4.3E-41** | -0.133 | 1.1E-05 | 0.309 | 5.7E-02 | -0.061 | 3.9E-01 | -0.087 | 2.9E-01 |
| *PTK2B* | -0.118 | 2.2E-01 | 0.005 | **6.9E-24** | 0.034 | 6.6E-03 | 0.392 | 4.5E-02 | -0.108 | 2.8E-01 | -0.136 | 8.2E-02 |
| *SLC24A4* | 0.032 | 1.5E-01 | -0.037 | **4.6E-19** | -0.087 | 4.0E-02 | -0.355 | 2.3E-01 | 0.036 | 4.0E-02 | -0.050 | 2.0E-01 |
| *SORL1* | 0.019 | 3.9E-01 | -0.215 | **3.3E-161** | -0.093 | **8.6E-09** | -0.202 | 1.7E-01 | 0.066 | 4.7E-02 | -0.062 | 3.9E-01 |
| *SPI1* | -0.173 | 4.5E-02 | -0.031 | **9.6E-08** | 0.004 | 8.6E-02 | 0.303 | 9.1E-02 | -0.005 | 7.4E-01 | 0.020 | 7.2E-01 |
| *TREM2* | 0.025 | 1.9E-01 | 0.001 | 8.9E-01 | 0.032 | 1.6E-01 | 0.139 | 3.3E-01 | NA | NA | NA | NA |
| *TSPOAP1* | -0.093 | 7.3E-02 | -0.017 | **1.4E-18** | -0.090 | 2.5E-03 | 0.360 | 5.1E-01 | -0.057 | 5.5E-02 | 0.125 | 7.3E-01 |
| *USP6NL* | 0.073 | 5.7E-01 | -0.020 | **1.6E-09** | 0.003 | 7.0E-02 | -0.501 | 1.8E-01 | 0.009 | 2.7E-01 | -0.179 | 2.9E-02 |
| *WWOX* | 0.040 | 9.6E-01 | -0.043 | **1.9E-15** | -0.076 | 1.0E-05 | -0.214 | 1.7E-01 | 0.062 | 2.4E-02 | 0.008 | 9.3E-01 |

Ast: astrocyte; ExN: excitatory neuron; InN: inhibitory neuron; Mic: microglia; Oli: oligodendrocytes; OPC: oligodendrocyte progenitor cells. Log2 fold change of expression levels (LogFC) and P value (P) were calculated between AD and control brains from single nuclei RNA sequencing data from ROSMAP. NA: not available due to small number of single cells. Significant false discovery rate (FDR) P values are highlighted in bold.

**Table S12.** Association of brain methylation level of the top ranked *APOE* ε2-related genes with neuropathological traits in the ROSMAP dataset

| Network.ID | Gene | CpG site | DistBP | Neuropath AD Diagnosis | | | Plaques | | | Tangles | | |
|---|---|---|---|---|---|---|---|---|---|---|---|---|
| | | | | β | SE | P-value | β | SE | P-value | β | SE | P-value |
| E2INT.1 | *ASAP1* | cg01466198 | 2137 | 13.93 | 7.92 | 7.9E-02 | -8.96 | 3.76 | 1.7E-02 | 9.97 | 3.73 | 7.8E-03 |
| E2INT.1 | *ECE1* | cg12460254 | -1181 | 2.73 | 2.51 | 2.8E-01 | -2.66 | 1.21 | 2.8E-02 | 0.66 | 1.21 | 5.9E-01 |
| E2INT.1; E2P.2 | *STAT5B* | cg23017654 | 9533 | 20.86 | 5.82 | **3.3E-04** | -6.09 | 2.60 | 1.9E-02 | 4.49 | 2.58 | 8.3E-02 |
| E2INT.2 | *DNAJC1* | cg06688642 | -3715 | 5.25 | 3.85 | 1.7E-01 | -3.97 | 1.84 | 3.1E-02 | 3.90 | 1.82 | 3.3E-02 |
| E2INT.2 | *EIPR1* | cg14074830 | 8063 | 6.00 | 9.51 | 5.3E-01 | -5.32 | 4.59 | 2.5E-01 | 9.21 | 4.55 | 4.3E-02 |
| E2P.1 | *ITGA1* | cg24935345 | 2526 | 5.85 | 2.31 | 1.1E-02 | -2.21 | 1.10 | 4.4E-02 | 0.45 | 1.09 | 6.8E-01 |
| E2P.1 | *PPP2CB* | cg23693487 | -2814 | -7.68 | 2.82 | 6.5E-03 | 4.48 | 1.32 | **7.3E-04** | -2.21 | 1.32 | 9.5E-02 |
| E2P.1 | *UBE2C* | cg24675094 | -2413 | 8.38 | 7.11 | 2.4E-01 | -4.19 | 3.35 | 2.1E-01 | 7.87 | 3.32 | 1.8E-02 |
| E2P.1; E2P.2 | *STAT3* | cg13467672 | -4197 | -9.36 | 4.00 | 1.9E-02 | 3.27 | 1.91 | 8.8E-02 | -2.99 | 1.90 | 1.2E-01 |
| E2P.2 | *CCL16* | cg14237025 | -525 | -7.76 | 3.91 | 4.7E-02 | 4.90 | 1.84 | 8.0E-03 | -2.46 | 1.84 | 1.8E-01 |
| E2P.2 | *CSGALNACT1* | cg08952343 | 7213 | 10.56 | 9.43 | 2.6E-01 | -11.06 | 4.48 | 1.4E-02 | 11.57 | 4.45 | 9.5E-03 |

CpG site: methylation array marker id; DistBP: distance in base pairs between the top-ranked CpG site and from the best SNP from *APOE* genotype stratified GWA results. CpG sites are within 10kb from the top ranked GWAS SNPs.
Significant P-values ($P<0.00135$) after multiple test correction are highlighted in bold.

**Table S13.** Association of levels of classical complement proteins with AD-related proteins

| Outcome | C1q | | | C4A | | | C4B | | | CRP | | |
|---|---|---|---|---|---|---|---|---|---|---|---|---|
| | β | SE | P-value | β | SE | P-value | β | SE | P-value | | SE | P-value |
| Aβ42 | -0.06 | 0.07 | 4.1E-01 | 0.02 | 0.07 | 7.9E-01 | -0.14 | 0.07 | 4.7E-02 | 0.07 | 0.07 | 3.3E-01 |
| pTau181 | 0.03 | 0.08 | 7.0E-01 | 0.17 | 0.08 | 3.4E-02 | -0.12 | 0.08 | 1.4E-01 | -0.02 | 0.08 | 8.4E-01 |
| pTau231 | -0.04 | 0.07 | 5.6E-01 | 0.16 | 0.07 | 2.3E-02 | -0.05 | 0.07 | 4.4E-01 | 0.20 | 0.07 | 2.4E-03 |
| tTau | 0.03 | 0.07 | 6.3E-01 | 0.00 | 0.07 | 9.9E-01 | 0.00 | 0.07 | 9.8E-01 | 0.01 | 0.07 | 9.2E-01 |
| pTau181/tTau | 0.15 | 0.08 | 6.0E-02 | 0.01 | 0.08 | 8.9E-01 | 0.09 | 0.08 | 2.6E-01 | 0.02 | 0.09 | 8.5E-01 |
| pTau231/tTau | 0.02 | 0.07 | 8.0E-01 | 0.12 | 0.07 | 8.2E-02 | 0.05 | 0.07 | 4.6E-01 | 0.13 | 0.07 | 4.3E-02 |
| C1q | NA | NA | NA | 0.08 | 0.08 | 3.3E-01 | 0.12 | 0.08 | 1.2E-01 | 0.22 | 0.08 | 4.4E-03 |
| C4A | 0.07 | 0.08 | 3.3E-01 | NA | NA | NA | 0.09 | 0.07 | 2.2E-01 | -0.01 | 0.07 | 8.8E-01 |
| C4B | 0.12 | 0.07 | 1.2E-01 | 0.09 | 0.07 | 2.2E-01 | NA | NA | NA | 0.19 | 0.07 | 9.8E-03 |
| CRP | 0.20 | 0.07 | 4.4E-03 | -0.01 | 0.07 | 8.8E-01 | 0.18 | 0.07 | 9.8E-03 | NA | NA | NA |

NA: not applicable.
